# Sensory profiling in classical Ehlers-Danlos syndrome: a case-control study revealing pain characteristics, somatosensory changes, and impaired pain modulation

**DOI:** 10.1101/2023.02.24.23286404

**Authors:** Marlies Colman, Delfien Syx, Inge de Wandele, Lies Rombaut, Deborah Wille, Zoë Malfait, Mira Meeus, Anne-Marie Malfait, Jessica Van Oosterwijck, Fransiska Malfait

## Abstract

Pain is one of the most important, yet poorly understood complaints in heritable connective tissue disorders (HCTD) caused by monogenic defects in extracellular matrix molecules. This is particularly the case for Ehlers-Danlos syndromes (EDS), paradigm collagen-related disorders. This study aimed to identify the pain signature and somatosensory characteristics in the rare classical type of EDS (cEDS) caused by defects in type V or rarely type I collagen. We used static and dynamic quantitative sensory testing and validated questionnaires in 19 individuals with cEDS and 19 matched controls.

Individuals with cEDS reported clinically relevant pain/discomfort (VAS ≥5/10 in 32% for average pain intensity the past month) and worse health -related quality of life. Altered sensory profile was found in the cEDS group with higher (p=0.04) detection thresholds for vibration stimuli at the lower limb indicating hypoesthesia, reduced thermal sensitivity with more (p<0.001) paradoxical thermal sensations, and hyperalgesia with lower pain thresholds to mechanical (p<0.001) stimuli at both the upper and lower limbs and to cold (p=0.005) stimulation at the lower limb. Using a parallel conditioned pain paradigm, the cEDS group showed significantly smaller antinociceptive responses (p-value between 0.005 and 0.046) suggestive of impaired endogenous central pain modulation.

In conclusion, Individuals with cEDS report chronic pain and worse health-related quality of life, and present altered somatosensory perception. This study is the first to systematically investigate pain and somatosensory characteristics in a genetically defined HCTD and provides interesting insights on the possible role of the ECM in the development and persistence of pain.

## 1. Introduction

The role of the extracellular matrix (ECM) in the development and persistence of pain has been increasingly recognized [80]. The ECM is a highly organized, dynamic network composed of structural (*e.g*., collagens and proteoglycans) and non-structural proteins, and is involved in many developmental, physiological and pathological processes [27]. Transcriptome analysis in mouse models of nerve injury- and inflammation-induced pain have identified ECM organization as an overrepresented molecular pathway [57], and functional and structural abnormalities of the nervous system and pain-related behaviors have been described in mouse models with genetic defects affecting the ECM [2; 3; 15; 78]. Strikingly, pain is highly prevalent in heritable connective tissue disorders (HCTD) caused by monogenic defects in ECM genes, including the Ehlers-Danlos syndromes (EDS) [45; 68; 86] and osteogenesis imperfecta [4], both collagen-related disorders, and Marfan syndrome [85], caused by defects in fibrillin-1. In fact, pain is the reason why many individuals with these conditions seek medical attention.

EDS is an umbrella term for a group of rare HCTD characterized by joint hypermobility, skin hyperextensibility, abnormal wound healing, easy bruising, and widespread connective tissue friability. Thirteen distinct EDS types are recognized with defects in 20 different genes that are involved in collagen biosynthesis, and/or supramolecular organization of collagen fibrils [44; 46]. With an estimated prevalence of 1:20,000, the autosomal dominant classical EDS type (cEDS; MIM #130000 and #130010) is the most common genetically elucidated EDS type [44]. Generalized joint hypermobility, skin hyperextensibility, skin fragility and atrophic scarring are the major clinical hallmarks of cEDS. Approximately 80-90% of the individuals with a clinical suspicion of cEDS harbor a defect in the *COL5A1* or *COL5A2* genes, which encode the proα1(V)- or proα2(V)-collagen chains of type V collagen, respectively [18]. Additionally, a rare arginine to cysteine substitution in the proα1(I)-collagen chain (*COL1A1*, c.934C>T, p.(Arg312Cys)) is found in a small fraction of cEDS patients [17].

Two questionnaire studies found self-reported chronic pain in >70% of individuals with cEDS [68; 86], but comprehensive data on pain characteristics and mechanisms in cEDS or other molecularly solved EDS types are currently non-existing. The few existing studies that have addressed pain in human EDS were conducted in heterogenous populations with hypermobile EDS (hEDS) or hypermobility spectrum disorders (HSD), which are molecularly unsolved and the diagnoses of which are based solely on clinical criteria [12; 46]. This major gap in the study of pain in EDS also hampers the development of effective treatment strategies for these individuals in whom the high use of analgesics, surgery, and physical therapy, brings only modest relief at best and is frequently associated with unwanted side effects [68; 86].

Interestingly, pain-related behavior and anormal cutaneous innervation were shown in a murine model of cEDS [78]. Hence, the current study aimed to investigate the somatosensory profile in human cEDS. The protocol included static and dynamic quantitative sensory testing (QST) with assessment of the sensitivity to different (non-)noxious stimulation modalities and evaluation of endogenous central pain modulation. Emotional and cognitive factors known to influence pain were assessed using validated questionnaires.

## 2. Materials and Methods

### 2.1 Study design and aims

The primary aim of this case-control study was to identify possible sensory alterations and study the mechanisms underlying chronic pain in individuals with cEDS. This study complied with the Declaration of Helsinki and was approved by the Ethical Committee of Ghent University Hospital (B670201941418). All participants were fully informed about the experimental procedures, and all provided written informed consent before inclusion. This study was reported according to the STrengthening the Reporting of OBservational studies in Epidemiology (STROBE) guideline for case-control studies [88].

### 2.2 Participants and guidelines

Individuals with molecularly confirmed cEDS were recruited from a previously reported cohort [18] (**Table 1**). Healthy age- and sex-matched pain-free controls were recruited among the hospital personnel, the region of the participating hospital and patients’ acquaintances and mutation-negative family members, the latter being a commonly applied method in pain research to minimize bias related to socio-economic status of participants. Dutch-speaking males and females between 18 and 65 years of age were eligible for study participation.

**Table 1:** characteristics and demographics of the study participants.

|  | **cEDS**<br>**n=19** | **Control**<br>**n=19** | **p-value** |
| --- | --- | --- | --- |
| **Sex** | 13F, 6M | 13F, 6M |  |
| **Age (years)** | 38.4 ± 13.1 | 36.6 ± 13.0 | 0.68 |
| **Weight (kg)** | 71.1 ± 16.4 | 68.8 ± 13.8 | 0.65 |
| **Height (cm)** | 170.3 ± 9.9 | 171.9 ± 8.9 | 0.62 |
| **BMI (kg/m<sup>2</sup>)** | 24.3 ± 4.1 | 23.24 ± 4.0 | 0.44 |
| **Blood pressure diastolic (mmHg)** | 88.7 ± 9.8 | 84.9 ± 10.3 | 0.25 |
| **Blood pressure systolic (mmHg)** | 123.9 ± 13.5 | 121.4 ± 10.8 | 0.53 |
| **Mean arterial pressure (mmHg)** | 100.5 (9.26) | 97.1 (2.15) | 0.27 |
| **Beighton score (0-9)** | 7 (5) | 0 (2) | ≤0.001 |
| **Skin hyperextensibility** | 19 | 1 | ≤0.0001 |
| **Atrophic scars** | 18 | 0 | ≤0.0001 |
| **Joint dislocations** | 13 | 0 | ≤0.0001 |
| **Subjective muscle weakness** | 9 | 0 | ≤0.001 |
| **Education** | High school: 7<br>Higher education: 7<br>University: 5 | High school: 7<br>Higher education: 5<br>University: 9 | 0.32 |
| **Smoker** | Yes: 1<br>Ex-smoker: 4 | Yes: 0<br>Ex-smoker: 2 | 0.38 |
| **Employement status** | Student: 2<br>Fulltime: 10<br>Parttime: 1<br>Retired: 2<br>Impairment: 4 | Student: 2<br>Fulltime: 14<br>Parttime: 1<br>Retired: 0<br>Impairment: 0 | 0.12 |

Normally distributed continuous variables are presented with 'mean ± standard deviation', non-normally distributed continuous variables as 'median (interquartile range)'.
BMI: Body Mass Index

Controls were excluded if they presented with generalized joint hypermobility (Beighton score >4/9), had a current pain problem or a history of chronic pain, or reported (daily) use of analgesics, anti-depressant, anxiolytic or antihypertensive medications. Additional exclusion criteria for both groups were the presence of any cardiovascular, respiratory, neurological or psychiatric conditions, pregnancy or breastfeeding in the past year or surgical interventions the past year.

### 2.3 Procedure

The study procedure is depicted in **Figure 1**. Before inclusion, all potential participants completed a pre-screening questionnaire assessing the eligibility criteria. All experimental procedures took place at the research laboratories of Ghent University/Ghent University Hospital. One week before the experimental procedures, all participants filled out a series of questionnaires using the secure web application RedCap [33]. These included a general questionnaire inquiring about the socio-demographics and medical history, the Central Sensitization Inventory (CSI) to assess self-reported symptoms related to central sensitization, the Hospital Anxiety and Depression Scale (HADS) to assess the presence of anxiety and depression, the International Physical Activity Questionnaire (IPAQ) to assess self-reported physical activity levels, and the Short-Form 36 health survey (SF-36) and the Health Assessment Questionnaire (HAQ) to assess health status and disability.

**Figure 1.**
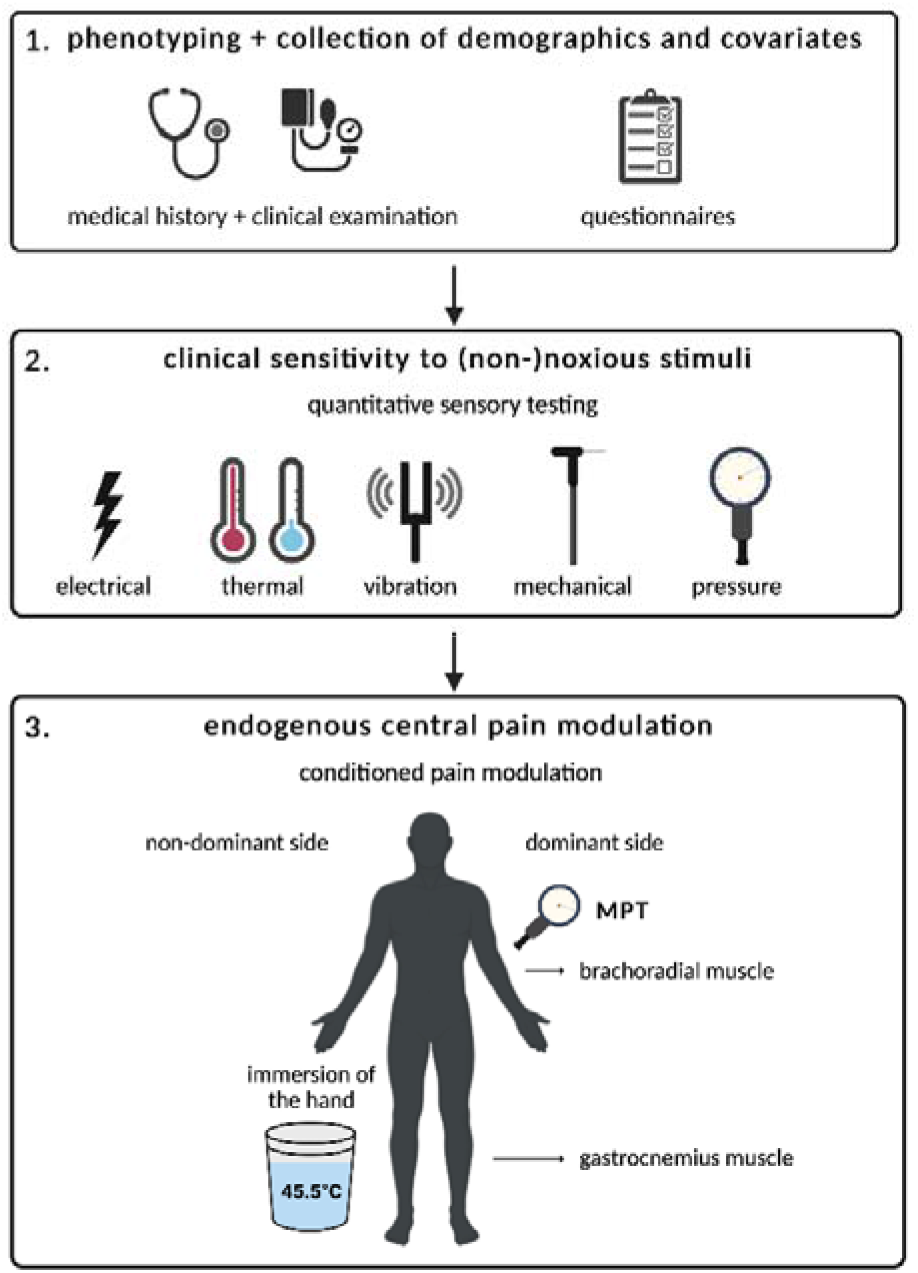
study procedure. The study protocol started with phenotyping of the participant and collection of demographics and covariates through self-report (questionnaires) and clinical examination (1). Next, static quantitative sensory testing (QST) was performed with assessment of clinical sensitivity to (non-)noxious electrical, thermal, vibration, and mechanical (touch and pressure) stimuli (2). The protocol was finished with dynamic QST which included parallel and sequential conditioned pain modulation (CPM) methods to assess endogenous central pain modulation (3). MPT: mechanical pain threshold

Participants were instructed to avoid intensive physical activity 24h prior to undergoing the experimental procedures, and to avoid intake of caffeine, nicotine, and alcohol 3 hours prior to the procedures. Only a light meal was allowed right before the experimental procedures. cEDS patients were instructed to refrain from opioid medication 24 hours prior to undergoing the experimental procedures and intake of short-acting analgesics and blood pressure agents was postponed until after the experimental procedures were finalized.

On the day of the experimental procedures, all participants first completed a set of questionnaires using RedCap to ensure 15-30 minutes of physical rest before starting the experimental procedures. They completed Visual Analogue Scales (VAS) to determine their actual pain intensity and their average pain intensity over the past 4 weeks, a Margolis body chart to pinpoint pain locations, the Pain Vigilance and Awareness Questionnaire (PVAQ) to assess the presence of pain hypervigilance and the Tampa Scale for Kinesiophobia (TSK) to assess fear of movement. The individuals with cEDS also filled out the PainDETECT questionnaire (PD-Q) and the Douleur Neuropathique 4 Questions questionnaire (DN4) to assess the possible presence of neuropathic pain.

Subsequently, clinical examination with assessment of blood pressure, weight, length, and clinical signs of connective tissue fragility (generalized joint hypermobility (Beighton score), skin hyperextensibility, presence of atrophic scars, bruising) was performed. This was followed by static QST which started with the assessment of electrical detection and pain thresholds, followed by thermal detection and pain thresholds (including paradoxical thermal sensations), vibration detection thresholds, and mechanical detection and pain thresholds. The protocol was concluded with dynamic QST consisting of a CPM paradigm. All experimental procedures took place in a sound-attenuated, temperature-controlled room (21-23°C) and were conducted by the same researcher (M.C.) using standardized instructions.

### 2.4 Self-reported measures

**Visual Analog Scales** (VAS) were used to measure current pain intensity, and average pain intensity over the past four weeks. A VAS is a continuous scale consisting of a 10 cm horizontal line with the left and right outer ends, respectively, labelled as no pain at all (score 0) and worst imaginable pain ever (score 10) [71]. The VAS has been shown to be a reliable and valid tool for the assessment of pain intensity [89].

Participants were asked to shade the areas on a **Margolis topographical body chart** that were painful for more than 24h the past 4 weeks and to highlight to most painful body area. The Margolis Pain Diagram uses two body outlines front and back, containing the 45 different areas [47].

The **Pain Detect Questionnaire (PD-Q)** is a validated self-reported screening tool for pain of neuropathic origin [28]. It comprises nine questions regarding the severity, course quality, and nature of the patient’s pain and specific neuropathic pain symptoms. The total score ranges from 0 to 38 and a score of >18 indicates that a predominantly neuropathic pain component is likely, whereas a total score ≤12 indicates that the pain is likely predominantly nociceptive. With a total score of 13– 18, the presence of neuropathic pain is ambiguous.

The **Douleur Neuropathique 4 Questions** (DN4) questionnaire aims to discriminate neuropathic pain from nociceptive pain with 10 items grouped into 4 sections [9]. The first 7 items inquire the quality of pain (burning, painful cold, electric shocks) and the presence of abnormal sensations (tingling, pins and needles, numbness, itching). The 3 remaining items are associated with a neurological examination of the painful area (touch hypoesthesia, pinprick hypoesthesia, tactile allodynia). A score of 1 is allocated to each positive item and a score of 0 to each negative item. A total score of ≥4/10 is used as a cut-off point for a possible neuropathic pain.

The **International Physical Activity Questionnaire** (IPAQ) estimates physical activity based on the reported activities during the last seven days [8]. Metabolic equivalents are calculated by multiplying the amount of minutes/week of physical activity with a factor that represents the strenuousness of the activities [19].

The **Health Assessment Questionnaire** (HAQ) measures self-reported activity limitations over the past 7 days using eight categories with different items: self-care, rising, eating, walking, hygiene, reach, grip and activities [29]. Each item is scored on a 4-point Likert scale from 0 (no difficulty) to 3 (unable). The highest component score in each category determines the score for the category. The eight category scores are averaged into an overall disability index. The disability index ranges from 0-3, where a score of 0-1 is interpreted as mild to moderate disability, 1-2 as moderate to severe disability and 2-3 as very severe disability [16].

The **Short-Form 36 health survey** (SF-36) measures quality-of-life and consists of 36 questions with standardized responses, organized into eight health domains: physical functioning, social functioning, role limitations due to physical problems, role limitations due to emotional problems, bodily pain, mental health, vitality and general health perception [90]. One additional item pertains to health change. All raw scores were linearly converted to a 0–100 scale providing sum scores for each domain. Lower scores indicate worse performance on the specific domain.

The **Central Sensitization Inventory** (CSI) is a self-reported questionnaire to assess the presence and severity of central sensitization in individuals with chronic pain [48]. It consists of 18 items that assess physical and psychological symptoms associated with central sensitization. Participants rate their symptoms on a 0 to 10 scale. Total scores range between 0-100 and higher scores indicate greater severity. Scores above 40 are defined as ‘central sensitization’.

The **Pain Vigilance and Awareness Questionnaire** (PVAQ) is a 16-item questionnaire to assess the attention, awareness, and vigilance to pain [49]. The items are scored on a 6-point Likert scale with 0 indicating ‘never’ and 5 indicating ‘continuously’. The total score ranges from 0 to 80 and a higher score is indicative of a higher degree of hypervigilance for pain.

The **TAMPA scale for kinesiophobia** (TSK) is a 17-item questionnaire that measures the fear of (re)injury due to movement [39]. The items are scored one a 4-point Likert scale and the total score ranges from 17 to 68, with higher scores corresponding to higher degrees of fear of movement. A total score >37 indicates high fear of movement [74].

The **Hospital Anxiety and Depression Scale** (HADS) is used to assess symptoms of anxiety and depression in individuals seeking medical treatment. It consists of 14 items, 7 of which assess symptoms of anxiety (anxiety subscale) and 7 of which assess symptoms of depression (depression subscale) [98]. Participants rate their symptoms on a 0 to 3 scale. Scores from each subscale range between 0 to 21, with a score of 8 or higher indicating the presence of anxiety and depression and score rages between 0-7 being normal, between 8-10 being mild, between 11-15 being moderate, and between 16-21 being severe [7].

The Dutch versions of the PD-Q, DN4, HADS, HAQ, SF-36, CSI, PVAQ, TSK have a good test–retest reliability, internal consistency and concurrent validity in populations with chronic pain conditions [1; 40; 62-64; 73; 75; 77; 81-84]. The Dutch IPAQ has been shown to be a reliable and reasonably valid physical activity measurement tool for the general adult population [84].

### 2.5 Static QST

**Electrical detection thresholds (EDT) and electrical pain thresholds (EPT)** were determined with transcutaneous electrical stimulation unilaterally at the lower limb using a bar electrode (Digitimer Ltd) placed over the sural nerve located in the retromalleolar path of the dominant leg (dermatome S1). The electrode was connected to a Digitimer DS7A constant current stimulator (Digitimer Ltd) [53].

The skin under the bar electrode was prepared by shaving (if necessary), scrubbing using Everi abrasive paste (Spes Medica) and degreasing with ether. The stimulus intensity was gradually increased and decreased using a method of limits starting at 2.0mA (train of 5 pulses at 250Hz). The stimulus intensity was decreased with steps of 0.5mA until the participant did not feel the stimulus anymore, the stimulus intensity was then increased with steps of 0.1mA until the participant felt the stimulus again, and this intensity was registered as the EDT. Subsequently, the stimulus intensity was further increased with steps of 0.5mA until the participant indicated the first feeling of discomfort and this intensity was registered as the EPT. The procedure was repeated three times. Participants were seated on a comfortable chair with 45°-55° knee flexion.

**Thermal detection and pain thresholds** were determined using a 2.5 × 5 cm TSA-II thermode (MEDOC TSA) connected to a Contact Heat-Evoked Potential Stimulator (CHEPS®, MEDOC) at the lower and the upper limb. More specifically, the stimuli were applied bilaterally at the proximal 1/3^rd^ of the calf and the brachioradial muscle. Once the participant indicated that the baseline temperature (32°C) of the thermode was perceived as thermoneutral on the skin, the temperature either increased or decreased by 1°C/s (limited between 0°C and 51°C for safety reasons) using a method of limits [65]. Participants were given a dual button switch to indicate the detection of temperature change and the threshold of discomfort (pain threshold). The participants were instructed to press the first button when they detected a change in temperature, which registered the corresponding temperature as the warm (WDT) or the cold detection threshold (CDT). When the participant first perceived the cold or heat stimuli as uncomfortable, they pressed the other button to register the corresponding temperature as the heat (HPT) or cold (CPT) pain threshold. At each location, six consecutive measurements were made (3 times warm, 3 times cold, in randomized order) and the participants were asked to indicate whether they felt warm or cold stimuli to determine paradoxical thermal sensations (PTS). In between each of the consecutive measurements, the thermode was slightly repositioned to avoid measurements at sensitized (preheated/precooled) skin. The participants were not able to watch the computer screen during the measurements. Assessments were performed with the participants in prone position.

**Vibration detection thresholds** (VDT) were determined bilaterally at lower and upper limb using a biothesiometer (Bio-Medical Instrument Co.). The tractor of the device was applied with uniform pressure on the medial malleoli and ulnar styloid processes. Participants were asked to inform the examiner of the first sensation of vibration as the amplitude of vibration was slowly increased by 1 V/s. The corresponding Hz was registered as the VDT. The measurement was repeated three times at each location by resetting the voltage to zero and again slowly increasing the voltage [66; 72]. The assessment was performed with the participants placed in supine position.

**Mechanical detection thresholds (MDT)** were measured bilaterally at the lower and upper limb, more specifically at the plantar side of the hallux and the middle of the hypothenar. MDT were determined using a standardized set of 20 Semmes-Weinstein Monofilaments (Touch Test Sensory Evaluator, Stoelting Co) with evaluator sizes between 1.65 and 6.65 (target force between 0.008 and 300 grams respectively). Using a method of limits, three threshold determinations were made, each with a series of descending (starting with evaluator size 5.07) and ascending stimulus intensities [65]. A skin contact time of about 2 seconds was ensured for each measurement [51; 65]. Participants were placed in supine position during the assessments.

**Mechanical pain thresholds (MPT)** were measured bilaterally at the lower and upper limb with an electronic pressure algometer with a 1 cm^2^ rubber disk tip (Force ten™ FDX, Wagner instruments). Three measurements were made at the proximal 1/3^rd^ of the calf and the brachioradial muscle. Pressure increased at a speed of 1kg/s, and participants were instructed to indicate the first feeling of discomfort. The corresponding kg/cm^2^ was registered as MPT [38; 65]. Assessments were performed with the participants in prone position.

For all final thresholds, the mean of three measurements at each test location was calculated and averaged. When performed bilaterally, measurements from both body sites were averaged. A variable interstimulus interval of 5-12s between consecutive assessments of the same measure was used.

### 2.6 Dynamic QST

**Conditioned pain modulation** (CPM) was evaluated using a of a heterotopic noxious conditioning stimulation protocol [96] during which the effect of a hot water immersion of the hand (i.e., conditioning stimulus) on pressure pain (i.e., test stimulus) was evaluated.

The conditioning stimulus was delivered by immersing the non-dominant hand in hot water which was circulated and heated to 45.5°C using a digital thermocouple heater (Polystat 36, Thermo Fisher Scientific). Previous research demonstrated a robust CPM effect during thermal stimulation at 45.5°C, while minimizing potential ceiling or floor effects [54; 93] and hot water immersion has been reported to have fair to excellent reliability [37] and provoke relatively large CPM effects [50]. During the protocol, the participant was able to see a timer to keep track of submersion time. If the participant was not able to complete six minutes of immersion, the duration of immersion was recorded. After two and six minutes of immersion, participants were asked to rate the pain intensity provoked by the hot water using a VAS-scale.

The test stimulus was determined as described in the previous section (MPT), this bilaterally prior to the conditioning stimulus (baseline) and two minutes after removal of the conditioning stimulus (sequential method). In addition, after the first 2 minutes of conditioning stimulus application the MPT were determined (parallel method) twice at the dominant side. The latter time point was selected based on findings of previous research indicating that CPM continues up to five minutes after removal of the conditioning stimulus [26] although it is typically observed that the magnitude of this after-effect does decrease over time. The use of pressure pain as test stimulus was previously shown to provide reliable CPM effects [36]. The participants were in prone position during the CPM protocol.

To evaluate the CPM effects, the absolute and relative differences between baseline MPTs and the parallel and sequential MPTs were calculated. A positive absolute change score (Δ) or a relative change score >1 indicates a decrease in the perceived pain intensity of the test stimulus and thus an increase of the MPT as a consequence of the conditioning stimulation and denotes a antinociceptive response [97]. A negative absolute change value or a relative change score <1 indicates an increase in the perceived pain intensity of the test stimulus and thus a decrease of the MPT as a consequence of the conditioning stimulation and denotes an pronociceptive response. Only the results of participants who completed the 6 min submersion were included in the CPM analyses based on the MPT calculation.

### 2.7 Statistical analysis

Statistical analysis was performed using IBM SPSS Statistics 28 (Statistical Package for the Social Sciences) (IBM SPSS Data Collection) and Graphpad Prism 9.4.0 (GraphPad Software). An *a priori* power analysis was performed for sample size estimation, based on data from a previous study regarding hyperalgesia in hEDS that compared MPT using pressure stimuli of 23 patients with hEDS to 23 healthy matched controls [67]. The effect sizes for the MPT at various test locations ranged from 1.11 to 1.36, which is considered large using Cohen’s criteria [30]. With an alpha of 0.05 and power of 0.95, the projected sample size to obtain a similar effect size for between-group comparisons ranges from n=16 to n=23 (G*Power 3.1.9.2) [22].

The normality of continuous variables was evaluated with Shapiro-Wilk tests, and inspection of histograms and QQ-plots. Comparisons between groups were performed with an independent Student’s t-test (normal distribution) or a Mann-Whitney U-test (deviation of normal distribution). Categorical variables were compared with a Chi^2^ test or Fisher’s Exact test. Results are expressed as mean ± 1 standard deviation (SD) or median with interquartile range (IQR), in case of a normal distribution or deviation of the normal distribution respectively for continuous variables, or as frequencies and percentages for categorical variables. The level of statistical significance was set to a p-value <0.05. To assess possible confounding, exploratory correlation analyses were performed but none reached statistical significance after the Holmes sequential Bonferroni correction. No adjustments were made for multiple comparisons as past research suggests that such corrections are considered to be overly conservative in cases in which outcome variables are correlated [59] as is the case among QST parameters [6].

## 3. Results

### 3.1 Study cohort

Nineteen cEDS patients (13 women, 6 men) and 19 healthy age- and sex-matched pain-free controls (13 women, 6 men) were recruited. The cEDS group (n=19) consisted of 17 individuals with a defect in *COL5A1*, one with a pathogenic *COL5A2* variant and one with the *COL1A1* c.934C>T, p.(Arg132Cys) variant. The specific pathogenic defects are summarized in **Sup. Table 1**. The cEDS and control group did not differ significantly regarding age, weight, height, body mass index (BMI), blood pressure (systolic, diastolic, mean arterial pressure), smoking, education level or employment status.

In the cEDS group, the severe connective tissue friability is reflected by some specific clinical characteristics (**Figure 2**). All individuals presented with a degree of skin hyperextensibility and all but one individual had atrophic scarring. Generalized joint hypermobility (Beighton score ≥5/9) was present in 14 cEDS patients (historical in 3 and absent in 2) while 13 experienced regular joint dislocations. Muscle weakness was reported by nine individuals. Eleven out of the 19 individuals with cEDS (58%) had ever taken analgesics (WHO step 1-2) for long existing musculoskeletal pain. Five cEDS patients (26%) took analgesics on a daily basis at the moment of the testing, with three taking medication within step 3 of the WHO-relief ladder. The characteristics of the cEDS group and control group are shown in **Table 1**, and clinical characteristics are shown in **Figure 2**.

**Figure 2.** clinical features (removed from preprint)

### 3.2 Self-reported measures

The results of the self-report questionnaires are summarized in **Table 2 and Sup. Fig. 1-2**. The VAS indicated that most individuals in the cEDS group experienced clinically relevant pain. On the day of the testing, the median VAS-score in the EDS group was 3/10 (IQR 2) with 21% (n=4) reporting a VAS-score of ≥5. When they were asked about pain experienced in the past four weeks, patients with cEDS reported a median VAS score of 7/10 (IQR 4) with a score of ≥5/10 in 68% (n=13) for the maximal pain intensity, and a median VAS score of 3/10 (IQR 3) with a score of ≥5/10 in 32% (n=6) for the average pain intensity.

**Table 2:** results of the questionnaires.

|  | cEDS<br>n=19 | Control<br>n=19 | p-value |
| --- | --- | --- | --- |
| **VAS current** | 3.0 (2) | 1 (0) | **≤0.001** |
| **VAS average pain last 4 weeks** | 3 (3) | 1 (1) | **≤0.001** |
| **VAS maximum pain last 4 weeks** | 7 (4) | 2 (2) | **≤0.001** |
| **PD-Q** | 9 (6) | / |  |
| **DN4** | 0 (2) | / |  |
| **CSI** | 41.2 ± 15.6 | 19.6 ± 8.4 | **≤0.001** |
| **HADS** Fear | 7.0 (7) | 4.00 (4) | **0.03** |
| Depression | 4 (6) | 1.00 (2) | **≤0.001** |
| **PVAQ** | 39.4 ± 11.0 | 27.5 ± 11.6 | **0.003** |
| **TSK** | 37.6 ± 5.0 | 24.9 ± 5.9 | **≤0.001** |
| **HAQ** | 0.1 (0.9) | 0 (0) | **≤0.001** |
| **IPAQ** | 2762.4 (3396.0) | 4618.03 (3382.5) | **0.015** |
| **SF-36** Physical functioning | 60 (40) | 100 (5) | **<0.001** |
| Limitations due to physical health | 75 (100) | 100 (0) | **<0.001** |
| Limitations due to emotional problems | 100 (33) | 100 (0) | 0.26 |
| Fatigue | 55 (25) | 70.00 (5) | **0.002** |
| Emotional well-being | 68.00 (28) | 80 (16) | **0.032** |
| Social functioning | 57.5 (45) | 90 (20) | **≤0.001** |
| Pain | 57.5 (43) | 90 (10) | **≤0.001** |
| General health | 48.7 ± 18.5 | 81.3 ± 14.0 | **≤0.001** |

Normally distributed continuous variables are presented with 'mean ± standard deviation', non-normally distributed continuous variables as 'median (Interquartile range)'.
VAS: Visual Analogue Scale, PD-Q: PainDetect Questionnaire, DN4: Douleur Neuropathique 4 Questions, CSI: Central Sensitization Inventory, HADS: Hospital Anxiety and Depression Scale, PVAQ: Pain Vigilance and Awareness Questionnaire, TSK: TAMPA scale for Kinesiophobia, HAQ: Health Assessment Questionnaire, IPAQ: International Physical Activity Questionnaire, SF-36: Short-Form 36 health survey

The localization of the pain on the body charts varied among the cEDS patients: in most cases pain was confined to a limited number of joints or lower back, although some also reported more generalized pain (**Sup. Fig. 3**). With only two cEDS (11%) scoring above the cut-off for possible neuropathic pain on the DN4 questionnaire and four cEDS individuals (21%) scoring above the cut-off for possible neuropathic pain on the PD-Q, there were no consistent self-reported neuropathic components to the pain in the cEDS group.

Physical activity (IPAQ) was significantly lower in the cEDS group compared to controls (p=0.015) and the HAQ scores were significantly higher in the cEDS group (p≤0.001) indicating moderate to severe impairment in 16% (n=3) of the individuals with cEDS. In addition, the cEDS group scored significantly worse on all subscales of the SF-36 (p-value between <0.001 and 0.032), except for the limitations due to emotional problems (p=0.26). The cEDS group also scored significantly higher on the CSI (p≤0.001) with the mean score (41.2 ± 15.6) above the cut-off (score >40) for central sensitization. Regarding cognitive-emotional factors, the cEDS group scored significantly higher on the PVAQ (p=0.003) and the TSK (p≤0.001) compared to the control group. With a mean score of 37.6 ± 5.0 on the TSK, the cEDS group scored above the cut-off (score>37) for high fear of movement. For the HADS, significantly higher scores were found in the cEDS group for both the fear (p=0.03) and depression (p≤0.001) subscales, but the median scores remained below the cut-off of mild-moderate symptoms of anxiety (median=7) or depression (median= 4).

### 3.3 Static QST

#### 3.3.1 Detection thresholds

No between-group differences were found for the EDT (p=0.15) and MDT (p=0.19 at the hypothenar and p=0.17 at the hallux), while a significantly higher VDT at the ankle (p=0.04) and a borderline significant VDT at the wrist (p=0.05) were noted in the cEDS group (**Figure 3A**).

No between-group differences were found for the thermal detection thresholds (CDT and WDT) at the brachioradialis (p=0.36) and the gastrocnemius (p=0.31) muscles. However, the cEDS group made significantly more mistakes (p≤0.001) in distinguishing heat and cold stimuli (PTS). In both groups, the majority of mistakes were made during measurements at the gastrocnemius muscle (72%), and most errors (60%) consisted of perceiving cold stimuli as warm (paradoxical heat) (**Figure 3B**).

**Figure 3:**
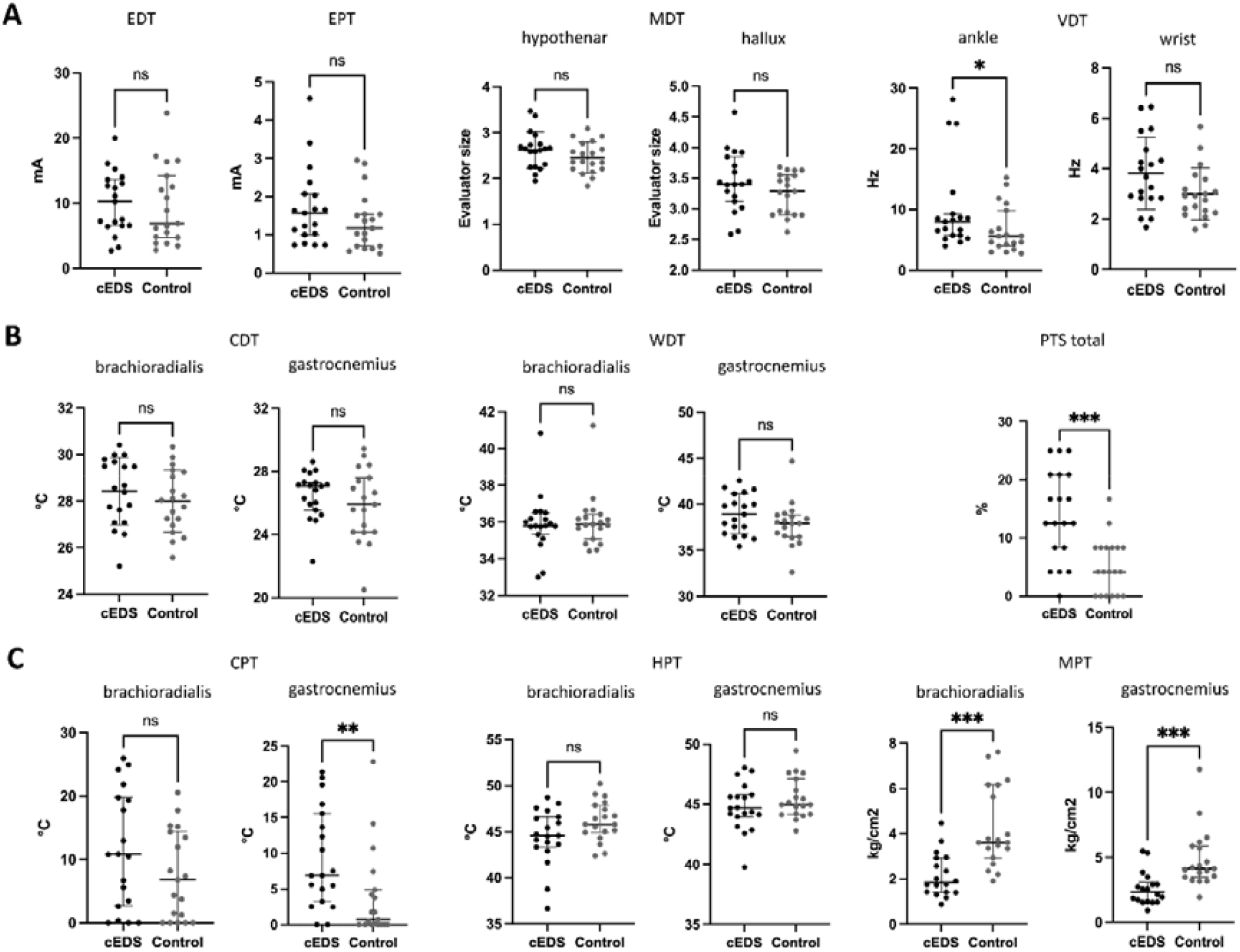
clinical sensitivity to innocuous and noxious stimuli. Normally distributed continuous variables are presented with ‘mean ± standard deviation, non-normally distributed continuous variables as ‘median (Interquartile range)’. EDT: electrical detection threshold, EPT: electrical pain threshold, MDT: mechanical detection threshold, VDT: vibration detection threshold, CDT: cold detection threshold, WDT: warm detection threshold, PTS: paradoxical thermal sensations, CPT: cold pain threshold, HPT: heat pain threshold, MPT: mechanical pain threshold, ns: non-significant, * p<0.05, ** p<0.01, *** p<0.001

#### 3.3.2 Pain thresholds

Regarding the pain thresholds, the EPT (p=0.54) and HPT (p=0.06 at the brachioradialis muscle and p=0.25 at the gastrocnemius muscle) did not differ significantly between both groups. For the CPT, the boundary of the measurement range (0°C) limited the determination of the CPT in both groups. However, a significantly higher CPT in the cEDS group (p=0.005) was found at the gastrocnemius muscle, while this was not the case at the brachioradialis muscle (p=0.21). The MPTs were significantly lower at both the brachioradialis (p≤0.001) and gastrocnemius (p≤0.001)) muscle in the cEDS group (**Figure 3C**).

Taken together, evidence was found for higher VDT, altered thermal sensitivity and increased sensitivity to painful mechanical and cold stimuli in the cEDS group. The results of the static QST are summarized in **Table 3**.

**Table 3:** quantitative sensory testing.

|  |  | **cEDS**<br>**n=19** | **Control**<br>**n=19** | **p-value** |
| --- | --- | --- | --- | --- |
| **EDT (mA)** |  | 1.57 (1.07) | 1.17 (0.83) | 0.15 |
| **EPT (mA)** |  | 10.23 (3.66) | 6.87 (9.47) | 0.54 |
| **CDT (°C)** | brachioradialis | 28.41 ± 1.44 | 28.00 ± 1.33 | 0.36 |
|  | gastrocnemius | 26.52 ± 1.49 | 25.88 ± 2.28 | 0.31 |
| **CPT (°C)** | brachioradialis | 10.80 (17.1) | 6.80 (14.38) | 0.21 |
|  | gastrocnemius | 6.88 (12.32) | 0.77 (4.89) | **0.005** |
| **WDT (°C)** | brachioradialis | 35.79 (1.17) | 35.88 (1.33) | 0.84 |
|  | gastrocnemius | 38.93 ± 2.19 | 37.83 ± 2.37 | 0.15 |
| **HPT (°C)** | brachioradialis | 44.46 ± 3.04 | 46.13 ± 2.15 | 0.06 |
|  | gastrocnemius | 44.80 ± 2.00 | 45.50 ± 1.71 | 0.25 |
| **PTS (%)** | total | 12.50 (12.50) | 4.17 (8.30) | **≤0.001** |
|  | brachioradialis | 0.00 (10.40) | 0.00 (8.30) | 0.84 |
|  | gastrocnemius | 25.00 (25) | 8.33 (8.30) | **≤0.001** |
| **VDT (Hz)** | wrist | 3.82 ± 1.43 | 3.00 ± 1.04 | **0.08** |
|  | ankle | 8.13 (4.40) | 5.58 (5.83) | **0.04** |
| **MDT (evaluator size)** | hypothenar | 2.62 ± 0.40 | 2.46 ± 0.34 | 0.19 |
|  | hallux | 3.44 ± 0.49 | 3.25 ± 0.34 | 0.17 |
| **MPT (kg/cm<sup>2</sup>)** | brachioradialis | 1.84 (1.52) | 3.61 (3.22) | **<0.001** |
|  | gastrocnemius | 2.35 (1.49) | 4.12 (2.38) | **<0.001** |

Normally distributed continuous variables are presented with 'mean ± standard deviation', non-normally distributed continuous variables as 'median (Interquartile range)'.
EDT: electrical detection threshold, EPT: electrical pain threshold, CDT: cold detection threshold, CPT: cold pain threshold, WDT: warm detection threshold, HPT: warm pain threshold, PTS: paradoxical thermal sensations, VDT: vibration detection threshold, MDT: mechanical detection threshold, MPT: mechanical pain threshold

### 3.4 Dynamic QST

Three individuals in the cEDS group (16%) and one in the control group (5%) did not finish the CPM protocol due to pain intolerance (VAS 10 within 2 minutes of the hot water immersion, p=0.6). On average, non-significant differences were found between the cEDS and control group regarding the VAS scores for the pain in the immersed hand given after 2 minutes (p=0.11) and at 6 minutes (p=0.08) of immersion. The MPTs remained significantly lower in the cEDS group compared to controls during the parallel and sequential protocols (p-values between ≤0.001 and 0.005) (**Sup. Table 2**).

During the parallel method, an antinociceptive response was found in 69% (n=11/16) at the brachioradial muscle and in 75% (n=12/16) at the gastrocnemius muscle of the cEDS group, and in 61% (n=11/18) at the brachioradial muscle and in 72% (n=13/18) at the gastrocnemius muscle of the control group. The sequential method showed an antinociceptive response at the brachioradial muscle in 94% (n=15/16) and at the gastrocnemius muscle in 81% (n=13/16) for the cEDS group. In the control group, this was the case in 67% (n=12/18) at both test locations. The absolute and relative CPM effects during the parallel and sequential protocols did not differ significantly between both groups (p-values between 1.00 and 0.31) (**Figure 4, Sup. Table 2**). However, when looking solely at the CPM effects of the individuals that showed antinociceptive responses, the CPM effects were significantly smaller in the cEDS group at both test locations for the parallel method, but not for the sequential method (**Sup. Fig. 4-5**).

**Figure 4:**
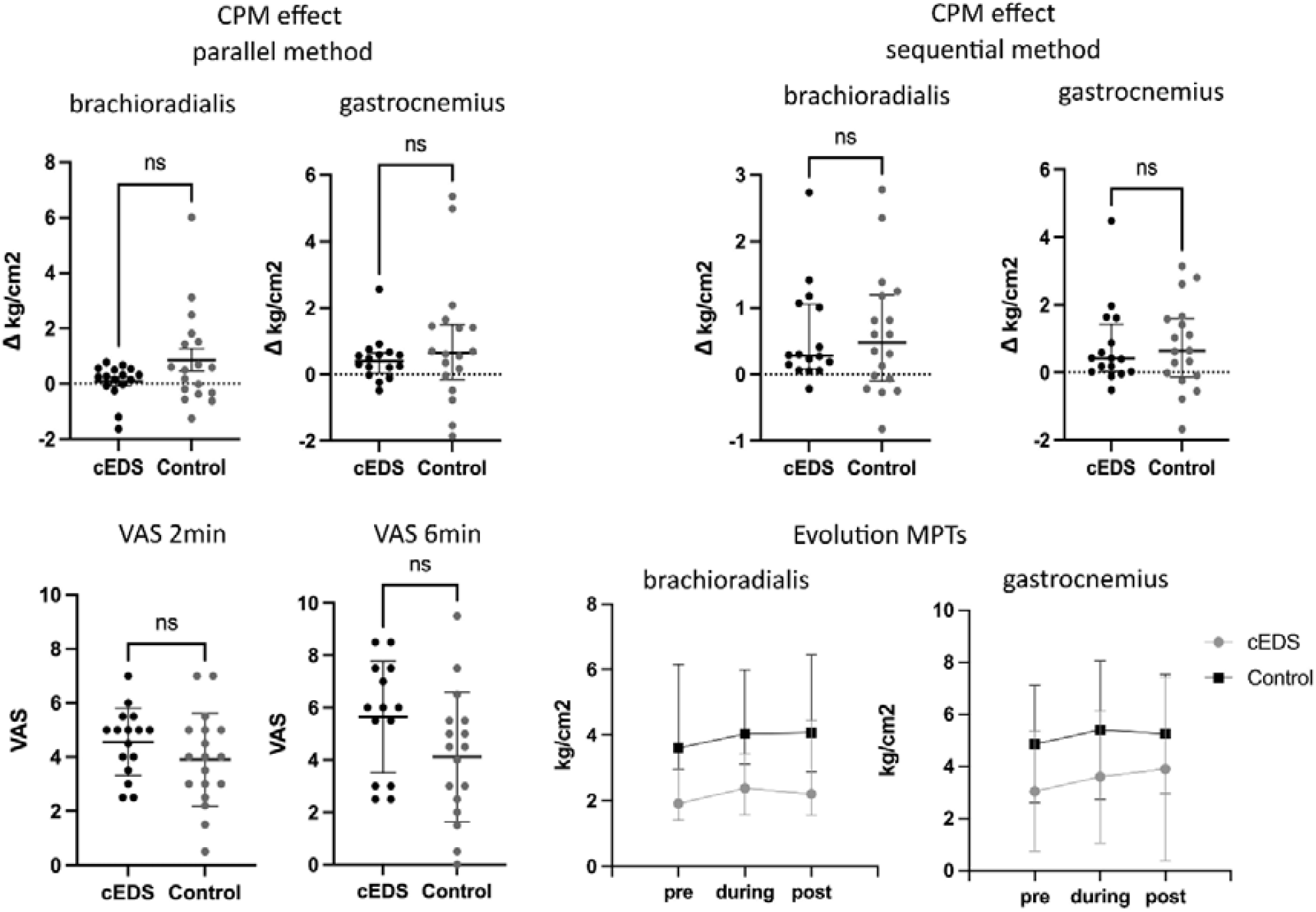
CPM effects. Absolute differences in MPT (kg/cm^2^) between the baseline measurement (pre) and the measurements during the parallel and sequential CPM methods. Normally distributed continuous variables are presented with ‘mean ± standard deviation, non-normally distributed continuous variables as ‘median (Interquartile range)’. MPT: mechanical pain threshold, CPM: conditioned pain modulation, VAS: visual analogue scale, ns: non-significant, ** p<0.01, *** p<0.001, **** p<0.0001.

## 4. Discussion

Pain is one of the major complaints in individuals with HCTD such as EDS, and existing treatment modalities are at best partially effective. To date, no studies have experimentally and quantitatively assessed somatosensory and pain mechanistic (dys)functions in genetically elucidated EDS types. To address this research gap in the study of pain associated with EDS, this study documented the somatosensory profile and pain signature of cEDS, using dynamic and static QST, and validated self-report measures.

The intensity and localization of the pain varied within the cEDS group. The majority experienced some chronic pain/discomfort, mostly at a limited number of joints or the spine, but more generalized pain as well as absence of any pain were also reported. The cEDS patients also experienced worse health-related quality of life with more limitations and impairment due to their disease. The results of this study further revealed changes in the somatosensory perception in cEDS, including signs of hypoesthesia with higher detection thresholds for vibration stimuli at the lower limb and reduced thermal sensitivity with more paradoxical thermal sensations at the lower limb, and signs of hyperalgesia with lower pain thresholds for cold stimuli at the lower limb and pressure stimuli at both the lower and upper limb. Moreover, the parallel CPM paradigm showed smaller antinociceptive responses in the cEDS group indicating impaired endogenous pain modulation. The results of the study are summarized **in Table 4**.

**Table 4:**
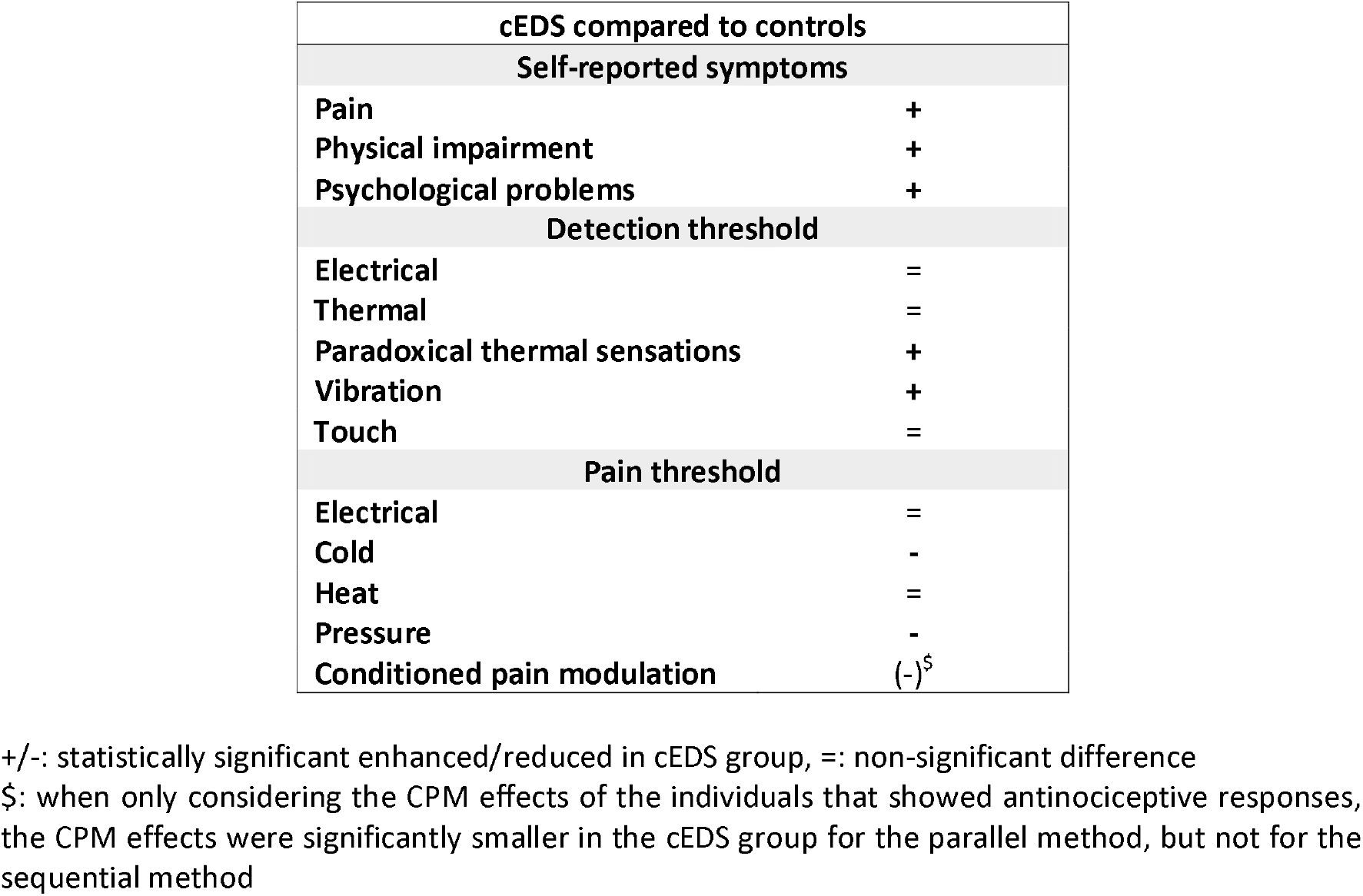
summary of the study results.

Mechanical and cold hyperalgesia, as found in the cEDS group, are believed to be mediated by sensitization of peripheral nociceptors (C-fibers, Aδ-fibers) [31]. Hyperalgesia to several stimulus types is also described in many neuropathic pain conditions [43] and chronic pain states with predominant nociplastic pain such as fibromyalgia [70] which is in the latter due to centrally mediated augmentation of pain facilitation or impaired pain inhibition, a process called central sensitization [94]. The cEDS group showed CSI scores above the cut-off score, indicative of central sensitization, but these scores need to be interpreted with caution as some items included in the questionnaire such as fatigue and gastro-intestinal complaints are also well-known symptoms of EDS [24; 45].

Vibration perception is mediated by both large (Aα) and medium diameter (Aβ) afferents which are also involved in proprioception [31], and diminished vibration perception as found in the cEDS group is also described in neuropathic pain conditions such as polyneuropathy [43]. Moreover, altered thermal sensibility with paradoxical heat is frequent in diverse neurological disorders including multiple sclerosis [32] and polyneuropathy [43] where impaired endogenous pain inhibition is caused by affection of inhibitory thalamic centers or malfunctioning Aδ-cold-fibers that disinhibit C-fiber nociceptors, respectively [76]. Impaired CPM has been described in several pronociceptive states, including central sensitization as well as neuropathic pain conditions [95].

Self-reported symptoms of neuropathic pain were not consistently present in the cEDS group. Interestingly, a questionnaire study reported neuropathic pain descriptors in 44% of individuals with Marfan Syndrome [52] while non-neuropathic pain was reported by adults with osteogenesis imperfecta [56]. In hEDS/HSD, a few questionnaire studies have reported a neuropathic component to hEDS/HSD-related pain [5; 11; 67; 87], although this is not consistently reported by all studies [21]. To add to this, decreased intra-epidermal nerve fiber density (IENFD) has been described in three case series of mainly hEDS/HSD patients. A first study reported decreased IENFD compared to normative values in 20 hEDS/HSD patients, three vascular EDS, and one cEDS patient [14; 58]. A second study reported abnormal IENFD compared to normative values in 78% of 69 individuals with hEDS/HSD [23], and a third report identified decreased IENFD in 61% of 31 hEDS patients compared to 16 healthy controls [35]. Additional reports investigating IENFD in individuals with genetically defined EDS types do not exist to date.

Previous work from our group in *Col5a1*^*+/-*^ mice, a well-studied mouse model of cEDS [91; 92], revealed a strikingly aberrant nociceptive innervation pattern of the glabrous skin in the footpad with a decreased number of nerves crossing the dermis-epidermis junction in combination with mechanical allodynia [78]. Pain-related behavior with increased sensitivity to chemical and mechanical, but not to thermal, stimuli, has also been demonstrated in *Tnxb*^*-/-*^ mice, a model for the autosomal recessive classical-like EDS (clEDS) caused by deficiency of Tenascin-X, a glycoprotein involved in the supramolecular organization of collagen fibrils [55]. A mouse model of osteogenesis imperfecta (*Col1a1*^*Jrt/+*^) showed hypersensitivity to mechanical and thermal stimuli but immunocytochemical analysis of the glabrous skin of the hind paw with the pan-neuronal marker PGP9.5 in revealed no changes in innervation [2].

As this is the first study to perform QST in cEDS, the results cannot be discussed in relation to previous findings and need to be confirmed by future studies. The findings of the experimental pain testing in this study partially differed from the existing studies in hEDS/HSD where inconsistent findings regarding somatosensory deficits have been reported. One report in hEDS/HSD found thermal and mechanical hypoesthesia [35] and one study described asymmetry in thermal and vibration detection thresholds when comparing the most painful joint and contralateral joint (no control group) [5], while no somatosensory deficits were found in other studies [20; 21; 35; 41; 66]. Cutaneous hyperesthesia [5; 13; 79] and hyperalgesia to pressure stimuli [20; 67; 69] are frequently found in hEDS/HSD. One study also found thermal hyperalgesia [21], but other reports identified no differences in thermal pain thresholds [20; 35; 41]. Evidence for increased pain facilitation and decreased pain inhibition was reported with increased wind-up for thermal and mechanical stimuli [5; 20; 21] and decreased exercise-induced analgesia [20], respectively. Conflicting findings, however, have been reported regarding central pain inhibition with CPM. One study found impaired CPM with contact heat as both test and conditioning stimulus [41], while no differences were found with repeated pressure stimuli as test stimulus and hot-water immersion as condition stimulus [20]. These different findings can possibly be attributed to differences in the used test protocols. Especially for the CPM paradigms, different protocols exist with different stimulus types, test locations, etc. While past recommendations advised the use of a sequential CPM method due to potentially less attention bias [97], a more recent study found no significant differences in CPM effects between sequential and parallel CPM methods [61]. Because of this lack of consensus, both a sequential and parallel CPM method were included in this study.

The burden of disease also impacted the psychological well-being of the cEDS group with mild/moderate symptoms of anxiety and depression, and more pain hypervigilance and kinesiophobia compared to healthy controls. This presence of mild to moderate emotional symptoms, but no severe symptoms or psychiatric conditions, is also reported in other HCTDs such as Marfan Syndrome [60] and osteogenesis imperfecta [25]. This contrasts with hEDS/HSD, in which psychiatric conditions including depression and anxiety disorders are more commonly observed [10; 34].

Taken together, our findings indicate an altered somatosensory function in cEDS and significantly add to the hypothesis that a general disturbance of the ECM can affect the organization and/or function of peripheral nerves, leading to changes in somatosensory perception. Furthermore, the previously reported structural alterations in dermal nociceptors of individuals with EDS and mouse models of EDS suggest that altered structural properties of skin nociceptors may contribute to the observed pain phenotype. More in depth and mechanistic investigations, such as assessment of small fiber architecture in individuals with cEDS and further clinical studies on proprioception are warranted to better understand the mechanisms underlying the observed changes in somatosensory perception in cEDS.

There are several limitations to this study. Although QST is commonly used to study neural (small fiber) function or pain sensitivity, it is prone to subjective bias (including attention, motivation etc.). This possible bias was minimized by using a single assessor and by using standardized instructions. Secondly, due to the rareness of cEDS, the sample size is limited which influences the power of this study. It will be critical to expand these studies to larger cohorts, highlighting the necessity of multicenter collaborations. The use of an age and sex matched control group in this study, on the other hand, is a major strength of this study as age is a known major determinant of sensory thresholds [42] and the inclusion of a matched control group is preferred above the use of reference values as the latter makes the study design more susceptible to confounding.

In conclusion, chronic pain is highly prevalent in cEDS and the disease has a profound influence on the individual’s quality of life and emotional well-being. Somatosensory profiling in cEDS showed presence of hyperalgesia for pressure and cold stimuli, hypoesthesia for vibration stimuli and alterations in thermal sensibility illustrated by the higher number of paradoxical thermal responses, and impaired endogenous pain modulation. This study is the first to investigate pain in a systematic way in a genetically defined group of individuals with a HCTD. Our findings indicate an altered neural function in cEDS providing interesting new insights on the possible role of the ECM in the development and persistence of pain.

## Data Availability

The data and program codes used in the analysis will not be made available to any researcher after publication. Subjects indicated that they had no objection of the use of anonymized data for this research, but they did not provide permission to use the data for other research.

## 5. Author contributions

**Marlies Colman**: conceptualization, study design, recruitment of participants, testing, processing of data, data analysis, interpretation of results, writing of the manuscript

**Delfien Syx**: recruitment of participants, interpretation of results, reviewing and editing of the manuscript

**Inge de Wandele**: conceptualization, study design, interpretation of results, reviewing and editing of the manuscript

**Lies Rombaut**: conceptualization, interpretation of results, reviewing and editing of the manuscript

**Deborah Wille**: recruitment of participants, assistance during testing, reviewing and editing of the manuscript

**Zoë Malfait**: assistance during testing, reviewing and editing of the manuscript

**Mira Meeus**: interpretation of results, reviewing and editing of the manuscript

**Anne-Marie Malfait**: conceptualization, reviewing and editing of the manuscript

**Jessica van Oosterwijck**: conceptualization, study design, reviewing and editing of the manuscript

**Fransiska Malfait**: conceptualization, study design, testing, reviewing and editing of the manuscript

## 7. Acknowledgements

We thank all participants included in this study. MC is a PhD research fellow funded by the Research Foundation − Flanders (FWO) [11B7921N]. DS is a postdoctoral fellow funded by the FWO [12Q5920N]. FM is a senior clinical investigator funded by the FWO [1842318N]. This work was supported by the FWO ([3G041519] to FM), Ghent University ([GOA019-21] to FM), Association française des syndromes d’Ehlers-Danlos (AFSED), The Ehlers-Danlos Society, Zebrapad vzw, and National Institute of Health, NIAMS, [P30AR079206] to AMM.

## References

[1] Aaronson NK, Muller M, Cohen PD, Essink-Bot M-L, Fekkes M, Sanderman R, Sprangers MA, Te Velde A, Verrips E. Translation, validation, and norming of the Dutch language version of the SF-36 Health Survey in community and chronic disease populations. Journal of clinical epidemiology 1998;51(11):1055–1068.

[2] Abdelaziz DM, Abdullah S, Magnussen C, Ribeiro-da-Silva A, Komarova SV, Rauch F, Stone LS. Behavioral signs of pain and functional impairment in a mouse model of osteogenesis imperfecta. Bone 2015;81:400–406.

[3] Aktar R, Peiris M, Fikree A, Cibert-Goton V, Walmsley M, Tough IR, Watanabe P, Araujo EJ, Mohammed SD, Delalande JM. The extracellular matrix glycoprotein tenascin-X regulates peripheral sensory and motor neurones. The Journal of Physiology 2018;596(17):4237–4251.

[4] Barlow S, Dove L, Jaggi A, Keen R, Bubbear J. The prevalence of musculoskeletal pain and therapy needs in adults with Osteogenesis Imperfecta (OI) a cross-sectional analysis. BMC Musculoskeletal Disorders 2022;23(1):1–7.

[5] Benistan K, Martinez V. Pain in hypermobile Ehlers-Danlos syndrome: New insights using new criteria. Am J Med Genet A 2019;179(7):1226–1234.

[6] Bhalang K, Sigurdsson A, Slade GD, Maixner W. Associations among four modalities of experimental pain in women. The Journal of Pain 2005;6(9):604–611.

[7] Bjelland I, Dahl AA, Haug TT, Neckelmann D. The validity of the Hospital Anxiety and Depression Scale: an updated literature review. Journal of psychosomatic research 2002;52(2):69–77.

[8] Booth M. Assessment of physical activity: an international perspective. Research quarterly for exercise and sport 2000;71(Sup2):114–120.

[9] Bouhassira D, Attal N, Alchaar H, Boureau F, Brochet B, Bruxelle J, Cunin G, Fermanian J, Ginies P, Grun-Overdyking A. Comparison of pain syndromes associated with nervous or somatic lesions and development of a new neuropathic pain diagnostic questionnaire (DN4). Pain 2005;114(1-2):29–36.

[10] Bulbena A, Baeza-Velasco C, Bulbena-Cabré A, Pailhez G, Critchley H, Chopra P, Mallorquí-Bagué N, Frank C, Porges S. Psychiatric and psychological aspects in the Ehlers–Danlos syndromes, Proceedings of the American Journal of Medical Genetics Part C: Seminars in Medical Genetics, Vol. 175: Wiley Online Library, 2017. pp. 237–245.

[11] Camerota F, Celletti C, Castori M, Grammatico P, Padua L. Neuropathic pain is a common feature in Ehlers-Danlos syndrome. J Pain Symptom Manage 2011;41(1):e2–4.

[12] Castori M, Hakim A. Contemporary approach to joint hypermobility and related disorders. Curr Opin Pediatr 2017;29(6):640–649.

[13] Castori M, Morlino S, Celletti C, Ghibellini G, Bruschini M, Grammatico P, Blundo C, Camerota F. Re-writing the natural history of pain and related symptoms in the joint hypermobility syndrome/Ehlers-Danlos syndrome, hypermobility type. Am J Med Genet A 2013;161a(12):2989–3004.

[14] Cazzato D, Castori M, Lombardi R, Caravello F, Bella ED, Petrucci A, Grammatico P, Dordoni C, Colombi M, Lauria G. Small fiber neuropathy is a common feature of Ehlers-Danlos syndromes. Neurology 2016;87(2):155–159.

[15] Chen P, Cescon M, Megighian A, Bonaldo P. Collagen VI regulates peripheral nerve myelination and function. Faseb j 2014;28(3):1145–1156.

[16] Cole JC, Motivala SJ, Khanna D, Lee JY, Paulus HE, Irwin MR. Validation of single-factor structure and scoring protocol for the Health Assessment Questionnaire-Disability Index. Arthritis Care & Research: Official Journal of the American College of Rheumatology 2005;53(4):536–542.

[17] Colman M, Castori M, Micale L, Ritelli M, Colombi M, Ghali N, Van Dijk F, Marsili L, Weeks A, Vandersteen A. Atypical variants in COL1A1 and COL3A1 associated with classical and vascular Ehlers-Danlos syndrome overlap phenotypes: expanding the clinical phenotype based on additional case reports. Clin Exp Rheumatol 2022;40:46–62.

[18] Colman M, Syx D, De Wandele I, Dhooge T, Symoens S, Malfait F. Clinical and molecular characteristics of 168 probands and 65 relatives with a clinical presentation of classical Ehlers-Danlos syndrome. Hum Mutat 2021;42(10):1294–1306.

[19] Craig CL, Marshall AL, Sjöström M, Bauman AE, Booth ML, Ainsworth BE, Pratt M, Ekelund U, Yngve A, Sallis JF. International physical activity questionnaire: 12-country reliability and validity. Medicine and science in sports and exercise 2003;35(8):1381–1395.

[20] De Wandele I, Colman M, Hermans L, Van Oosterwijck J, Meeus M, Descheemaeker F, Calders P, Rombaut L, Brusselmans G, Syx D, De Paepe A, Malfait F. Pain modulation in patients with hypermobile EDS and hypermobility spectrum disorders - a pilot study. 2020.

[21] Di Stefano G, Celletti C, Baron R, Castori M, Di Franco M, La Cesa S, Leone C, Pepe A, Cruccu G, Truini A, Camerota F. Central sensitization as the mechanism underlying pain in joint hypermobility syndrome/Ehlers-Danlos syndrome, hypermobility type. Eur J Pain 2016;20(8):1319–1325.

[22] Faul F, Erdfelder E, Lang A-G, Buchner A. G* Power 3: A flexible statistical power analysis program for the social, behavioral, and biomedical sciences. Behavior research methods 2007;39(2):175–191.

[23] Fernandez A, Aubry-Rozier B, Vautey M, Berna C, Suter MR. Small fiber neuropathy in hypermobile Ehlers Danlos syndrome/hypermobility spectrum disorder. J Intern Med 2022;292(6):957.

[24] Fikree A, Chelimsky G, Collins H, Kovacic K, Aziz Q. Gastrointestinal involvement in the Ehlers–Danlos syndromes, Proceedings of the American Journal of Medical Genetics Part C: Seminars in Medical Genetics, Vol. 175: Wiley Online Library, 2017. pp. 181–187.

[25] Forestier-Zhang L, Watts L, Turner A, Teare H, Kaye J, Barrett J, Cooper C, Eastell R, Wordsworth P, Javaid MK. Health-related quality of life and a cost-utility simulation of adults in the UK with osteogenesis imperfecta, X-linked hypophosphatemia and fibrous dysplasia. Orphanet J Rare Dis 2016;11(1):1–9.

[26] France CR, Suchowiecki S. A comparison of diffuse noxious inhibitory controls in men and women. Pain 1999;81(1-2):77–84.

[27] Frantz C, Stewart KM, Weaver VM. The extracellular matrix at a glance. J Cell Sci 2010;123(24):4195–4200.

[28] Freynhagen R, Baron R, Gockel U, Tölle TR. Pain DETECT: a new screening questionnaire to identify neuropathic components in patients with back pain. Curr Med Res Opin 2006;22(10):1911–1920.

[29] Fries JF, Spitz P, Kraines RG, Holman HR. Measurement of patient outcome in arthritis. Arthritis & Rheumatism 1980;23(2):137–145.

[30] Fritz CO, Morris PE, Richler JJ. Effect size estimates: current use, calculations, and interpretation. Journal of experimental psychology: General 2012;141(1):2.

[31] Goetz CG. Textbook of clinical neurology, Vol. 355: Elsevier Health Sciences, 2007.

[32] Hansen C, Hopf H, Treede R. Paradoxical heat sensation in patients with multiple sclerosis: evidence for a supraspinal integration of temperature sensation. Brain 1996;119(5):1729–1736.

[33] Harris PA, Taylor R, Thielke R, Payne J, Gonzalez N, Conde JG. Research electronic data capture (REDCap)—A metadata-driven methodology and workflow process for providing translational research informatics support. Journal of Biomedical Informatics 2009;42(2):377–381.

[34] Hershenfeld SA, Wasim S, McNiven V, Parikh M, Majewski P, Faghfoury H, So J. Psychiatric disorders in Ehlers–Danlos syndrome are frequent, diverse and strongly associated with pain. Rheumatology International 2016;36(3):341–348.

[35] Igharo D, Thiel JC, Rolke R, Akkaya M, Weis J, Katona I, Schulz JB, Maier A. Skin biopsy reveals generalized small fibre neuropathy in hypermobile Ehlers–Danlos syndromes. Eur J Neurol 2022.

[36] Imai Y, Petersen KK, Mørch CD, Arendt Nielsen L. Comparing test–retest reliability and magnitude of conditioned pain modulation using different combinations of test and conditioning stimuli. Somatosensory & motor research 2016;33(3-4):169–177.

[37] Kennedy DL, Kemp HI, Ridout D, Yarnitsky D, Rice AS. Reliability of conditioned pain modulation: a systematic review. Pain 2016;157(11):2410.

[38] Kinser AM, Sands WA, Stone MH. Reliability and validity of a pressure algometer. The Journal of Strength & Conditioning Research 2009;23(1):312–314.

[39] Kori S. Kinesiophobia: a new view of chronic pain behavior. Pain Manage 1990;3:35–43.

[40] Kregel J, Vuijk PJ, Descheemaeker F, Keizer D, van der Noord R, Nijs J, Cagnie B, Meeus M, van Wilgen P. The Dutch Central Sensitization Inventory (CSI): factor analysis, discriminative power, and test-retest reliability. The Clinical journal of pain 2016;32(7):624–630.

[41] Leone CM, Celletti C, Gaudiano G, Puglisi PA, Fasolino A, Cruccu G, Camerota F, Truini A. Pain due to Ehlers-Danlos Syndrome Is Associated with Deficit of the Endogenous Pain Inhibitory Control. Pain medicine (Malden, Mass) 2020;21(9):1929–1935.

[42] Lin YH, Hsieh SC, Chao CC, Chang YC, Hsieh ST. Influence of aging on thermal and vibratory thresholds of quantitative sensory testing. Journal of the Peripheral Nervous System 2005;10(3):269–281.

[43] Maier C, Baron R, Tölle T, Binder A, Birbaumer N, Birklein F, Gierthmühlen J, Flor H, Geber C, Huge V. Quantitative sensory testing in the German Research Network on Neuropathic Pain (DFNS): somatosensory abnormalities in 1236 patients with different neuropathic pain syndromes. Pain 2010;150(3):439–450.

[44] Malfait F, Castori M, Francomano CA, Giunta C, Kosho T, Byers PH. The Ehlers–Danlos syndromes. Nature Reviews Disease Primers 2020;6(1):64.

[45] Malfait F, Colman M, Vroman R, De Wandele I, Rombaut L, Miller RE, Malfait AM, Syx D. Pain in the Ehlers-Danlos syndromes: Mechanisms, models, and challenges. Am J Med Genet C Semin Med Genet 2021;187(4):429–445.

[46] Malfait F, Francomano C, Byers P, Belmont J, Berglund B, Black J, Bloom L, Bowen JM, Brady AF, Burrows NP, Castori M, Cohen H, Colombi M, Demirdas S, De Backer J, De Paepe A, Fournel-Gigleux S, Frank M, Ghali N, Giunta C, Grahame R, Hakim A, Jeunemaitre X, Johnson D, Juul-Kristensen B, Kapferer-Seebacher I, Kazkaz H, Kosho T, Lavallee ME, Levy H, Mendoza-Londono R, Pepin M, Pope FM, Reinstein E, Robert L, Rohrbach M, Sanders L, Sobey GJ, Van Damme T, Vandersteen A, van Mourik C, Voermans N, Wheeldon N, Zschocke J, Tinkle B. The 2017 international classification of the Ehlers-Danlos syndromes. Am J Med Genet C Semin Med Genet 2017;175(1):8–26.

[47] Margolis RB, Tait RC, Krause SJ. A rating system for use with patient pain drawings. Pain 1986;24(1):57–65.

[48] Mayer TG, Neblett R, Cohen H, Howard KJ, Choi YH, Williams MJ, Perez Y, Gatchel RJ. The development and psychometric validation of the central sensitization inventory. Pain Practice 2012;12(4):276–285.

[49] McCracken LM. “Attention” to pain in persons with chronic pain: A behavioral approach. Behavior therapy 1997;28(2):271–284.

[50] Mertens MG, Hermans L, Crombez G, Goudman L, Calders P, Van Oosterwijck J, Meeus M. Comparison of five conditioned pain modulation paradigms and influencing personal factors in healthy adults. European Journal of Pain 2021;25(1):243–256.

[51] Mücke M, Cuhls H, Radbruch L, Baron R, Maier C, Tölle T, Treede R-D, Rolke R. Quantitative sensory testing (QST). English version. Der Schmerz 2021;35(3):153–160.

[52] Nelson AM, Walega DR, McCarthy RJ. The incidence and severity of physical pain symptoms in Marfan syndrome. The Clinical journal of pain 2015;31(12):1080–1086.

[53] Neziri AY, Curatolo M, Nüesch E, Scaramozzino P, Andersen OK, Arendt-Nielsen L, Jüni P. Factor analysis of responses to thermal, electrical, and mechanical painful stimuli supports the importance of multi-modal pain assessment. Pain 2011;152(5):1146–1155.

[54] Nir R-R, Yarnitsky D, Honigman L, Granot M. Cognitive manipulation targeted at decreasing the conditioning pain perception reduces the efficacy of conditioned pain modulation. Pain 2012;153(1):170–176.

[55] Okuda-Ashitaka E, Kakuchi Y, Kakumoto H, Yamanishi S, Kamada H, Yoshidu T, Matsukawa S, Ogura N, Uto S, Minami T. Mechanical allodynia in mice with tenascin-X deficiency associated with Ehlers-Danlos syndrome. Sci Rep 2020;10(1):1–12.

[56] Orlando G, Pinedo-Villanueva R, Reeves ND, Javaid MK, Ireland A. Physical function in UK adults with osteogenesis imperfecta: a cross-sectional analysis of the RUDY study. Osteoporosis International 2021;32(1):157–164.

[57] Parisien M, Samoshkin A, Tansley SN, Piltonen MH, Martin LJ, El-Hachem N, Dagostino C, Allegri M, Mogil JS, Khoutorsky A, Diatchenko L. Genetic pathway analysis reveals a major role for extracellular matrix organization in inflammatory and neuropathic pain. Pain 2019;160(4):932–944.

[58] Pascarella A, Provitera V, Lullo F, Stancanelli A, Saltalamacchia AM, Caporaso G, Nolano M. Evidence of small fiber neuropathy in a patient with Ehlers-Danlos syndrome, hypermobility-type. Clinical neurophysiology : official journal of the International Federation of Clinical Neurophysiology 2016;127(3):1914–1916.

[59] Pocock SJ. Clinical trials: a practical approach: John Wiley & Sons, 2013.

[60] Rand-Hendriksen S, S⊘rensen I, Holmström H, Andersson S, Finset A. Fatigue, cognitive functioning and psychological distress in Marfan syndrome, a pilot study. Psychology, health & medicine 2007;12(3):305–313.

[61] Reezigt RR, Kielstra SC, Coppieters MW, Scholten-Peeters GG. No relevant differences in conditioned pain modulation effects between parallel and sequential test design. A cross-sectional observational study. PeerJ 2021;9:e12330.

[62] Roelofs J, Goubert L, Peters ML, Vlaeyen JW, Crombez G. The Tampa Scale for Kinesiophobia: further examination of psychometric properties in patients with chronic low back pain and fibromyalgia. European Journal of Pain 2004;8(5):495–502.

[63] Roelofs J, Peters M, Muris P, Vlaeyen J. Dutch version of the Pain Vigilance and Awareness Questionnaire: validity and reliability in a pain-free population. Behaviour Research and Therapy 2002;40(9):1081–1090.

[64] Roelofs J, Peters ML, McCracken L, Vlaeyen JW. The pain vigilance and awareness questionnaire (PVAQ): further psychometric evaluation in fibromyalgia and other chronic pain syndromes. Pain 2003;101(3):299–306.

[65] Rolke R, Baron R, Maier C, Tolle TR, Treede RD, Beyer A, Binder A, Birbaumer N, Birklein F, Botefur IC, Braune S, Flor H, Huge V, Klug R, Landwehrmeyer GB, Magerl W, Maihofner C, Rolko C, Schaub C, Scherens A, Sprenger T, Valet M, Wasserka B. Quantitative sensory testing in the German Research Network on Neuropathic Pain (DFNS): standardized protocol and reference values. Pain 2006;123(3):231–243.

[66] Rombaut L, De Paepe A, Malfait F, Cools A, Calders P. Joint position sense and vibratory perception sense in patients with Ehlers-Danlos syndrome type III (hypermobility type). Clin Rheumatol 2010;29(3):289–295.

[67] Rombaut L, Scheper M, De Wandele I, De Vries J, Meeus M, Malfait F, Engelbert R, Calders P. Chronic pain in patients with the hypermobility type of Ehlers-Danlos syndrome: evidence for generalized hyperalgesia. Clin Rheumatol 2015;34(6):1121–1129.

[68] Sacheti A, Szemere J, Bernstein B, Tafas T, Schechter N, Tsipouras P. Chronic pain is a manifestation of the Ehlers-Danlos syndrome. J Pain Symptom Manage 1997;14(2):88–93.

[69] Scheper MC, Pacey V, Rombaut L, Adams RD, Tofts L, Calders P, Nicholson LL, Engelbert RH. Generalized Hyperalgesia in Children and Adults Diagnosed With Hypermobility Syndrome and Ehlers-Danlos Syndrome Hypermobility Type: A Discriminative Analysis. Arthritis Care Res (Hoboken) 2017;69(3):421–429.

[70] Schmidt-Wilcke T, Clauw DJ. Fibromyalgia: from pathophysiology to therapy. Nature Reviews Rheumatology 2011;7(9):518–527.

[71] Scott J, Huskisson E. Graphic representation of pain. Pain 1976;2(2):175–184.

[72] Shakoor N, Agrawal A, Block JA. Reduced lower extremity vibratory perception in osteoarthritis of the knee. Arthritis Care & Research: Official Journal of the American College of Rheumatology 2008;59(1):117–121.

[73] Siegert CE, Vleming LJ, Vandenbroucke JP, Cats A. Measurement of disability in Dutch rheumatoid arthritis patients. Clin Rheumatol 1984;3(3):305–309.

[74] Silver A, Haeney M, Vijayadurai P, Wilks D, Pattrick M, Main C. The role of fear of physical movement and activity in chronic fatigue syndrome. Journal of psychosomatic research 2002;52(6):485–493.

[75] Spinhoven P, Ormel J, Sloekers P, Kempen G, Speckens AE, van Hemert AM. A validation study of the Hospital Anxiety and Depression Scale (HADS) in different groups of Dutch subjects. Psychological medicine 1997;27(2):363–370.

[76] Susser E, Sprecher E, Yarnitsky D. Paradoxical heat sensation in healthy subjects: peripherally conducted by Aδ or C fibres? Brain 1999;122(2):239–246.

[77] Swinkels-Meewisse E, Swinkels R, Verbeek A, Vlaeyen J, Oostendorp R. Psychometric properties of the Tampa Scale for kinesiophobia and the fear-avoidance beliefs questionnaire in acute low back pain. Man Ther 2003;8(1):29–36.

[78] Syx D, Miller RE, Obeidat AM, Tran PB, Malfait Z, Miller RJ, Malfait F, Malfait AM. Pain-related behaviors and abnormal cutneous innervation in a murine model of classical Ehlers-Danlos syndrome. Pain (03043959) 2020.

[79] Syx D, Symoens S, Steyaert W, De Paepe A, Coucke PJ, Malfait F. Ehlers-Danlos Syndrome, Hypermobility Type, Is Linked to Chromosome 8p22-8p21.1 in an Extended Belgian Family. Dis Markers 2015;2015:828970.

[80] Tajerian M, Clark JD. The role of the extracellular matrix in chronic pain following injury. Pain 2015;156(3):366–370.

[81] ten Klooster PM, Taal E, Van De Laar MA. Rasch analysis of the Dutch Health Assessment Questionnaire disability index and the Health Assessment Questionnaire II in patients with rheumatoid arthritis. Arthritis Care Res (Hoboken) 2008;59(12):1721–1728.

[82] Timmerman H, Wolff AP, Schreyer T, Outermans J, Evers AW, Freynhagen R, Wilder-Smith OH, van Zundert J, Vissers KC. Cross-cultural adaptation to the Dutch language of the PainDETECT-Questionnaire. Pain Practice 2013;13(3):206–214.

[83] van Seventer R, Vos C, Giezeman M, Meerding WJ, Arnould B, Regnault A, van Eerd M, Martin C, Huygen F. Validation of the Dutch version of the DN 4 diagnostic questionnaire for neuropathic pain. Pain Practice 2013;13(5):390–398.

[84] Vandelanotte C, De Bourdeaudhuij I, Philippaerts R, Sjöström M, Sallis J. Reliability and validity of a computerized and Dutch version of the International Physical Activity Questionnaire (IPAQ). J Phys Act Health 2005;2(1):63–75.

[85] Velvin G, Bathen T, Rand-Hendriksen S, Geirdal A. Systematic review of chronic pain in persons with Marfan syndrome. Clin Genet 2016;89(6):647–658.

[86] Voermans NC, Knoop H, Bleijenberg G, van Engelen BG. Pain in ehlers-danlos syndrome is common, severe, and associated with functional impairment. J Pain Symptom Manage 2010;40(3):370–378.

[87] Voermans NC, Knoop H, van Engelen BG. High frequency of neuropathic pain in Ehlers-Danlos syndrome: an association with axonal polyneuropathy and compression neuropathy? J Pain Symptom Manage 2011;41(5):e4–6; author reply e6-7.

[88] Von Elm E, Altman DG, Egger M, Pocock SJ, Gøtzsche PC, Vandenbroucke JP. The Strengthening the Reporting of Observational Studies in Epidemiology (STROBE) statement: guidelines for reporting observational studies. The Lancet 2007;370(9596):1453–1457.

[89] Von Korff M, Jensen MP, Karoly P. Assessing global pain severity by self-report in clinical and health services research. Spine 2000;25(24):3140–3151.

[90] Ware Jr JE, Sherbourne CD. The MOS 36-item short-form health survey (SF-36): I. Conceptual framework and item selection. Medical care 1992:473–483.

[91] Wenstrup RJ, Florer JB, Davidson JM, Phillips CL, Pfeiffer BJ, Menezes DW, Chervoneva I, Birk DE. Murine model of the Ehlers-Danlos syndrome. col5a1 haploinsufficiency disrupts collagen fibril assembly at multiple stages. J Biol Chem 2006;281(18):12888–12895.

[92] Wenstrup RJ, Smith SM, Florer JB, Zhang G, Beason DP, Seegmiller RE, Soslowsky LJ, Birk DE. Regulation of collagen fibril nucleation and initial fibril assembly involves coordinate interactions with collagens V and XI in developing tendon. J Biol Chem 2011;286(23):20455–20465.

[93] Wicksell RK, Olsson GL, Hayes SC. Psychological flexibility as a mediator of improvement in Acceptance and Commitment Therapy for patients with chronic pain following whiplash. European journal of pain 2010;14(10):1059. e1051-1059. e1011.

[94] Woolf CJ. Central sensitization: implications for the diagnosis and treatment of pain. Pain 2011;152(3):S2–S15.

[95] Yarnitsky D. Conditioned pain modulation (the diffuse noxious inhibitory control-like effect): its relevance for acute and chronic pain states. Current Opinion in Anesthesiology 2010;23(5):611–615.

[96] Yarnitsky D, Arendt-Nielsen L, Bouhassira D, Edwards RR, Fillingim RB, Granot M, Hansson P, Lautenbacher S, Marchand S, Wilder-Smith O. Recommendations on terminology and practice of psychophysical DNIC testing. European Journal of Pain-Kidlington 2010;14(4):339.

[97] Yarnitsky D, Bouhassira D, Drewes A, Fillingim R, Granot M, Hansson P, Landau R, Marchand S, Matre D, Nilsen K. Recommendations on practice of conditioned pain modulation (CPM) testing. European journal of pain 2015;19(6):805–806.

[98] Zigmond AS, Snaith RP. The hospital anxiety and depression scale. Acta psychiatrica scandinavica 1983;67(6):361–370.

