## Supplementary information for "Sensory profiling in classical Ehlers-Danlos syndrome: a case-control study revealing pain characteristics, somatosensory changes, and impaired pain modulation"

### Sensory profiling in monogenic alterations of the extracellular matrix: evidence for altered neural function in classical Ehlers-Danlos syndrome.

Marlies Colman^1,2,4^, Delfien Syx^1,2^, Inge de Wandele^2^, Lies Rombaut^2^, Deborah Wille^2^, Zoë Malfait^2^, Mira Meeus^3,4^, Anne-Marie Malfait^5^, Jessica Van Oosterwijck^3,4^, Fransiska Malfait^1,2^

1. Center for Medical Genetics, Ghent University Hospital, Ghent, Belgium

2. Department of Biomolecular Medicine, Faculty of Medicine and Health Sciences, Ghent University, Ghent, Belgium

3. Spine, Head and Pain SPINE Research Unit Ghent, Faculty of Medicine and Health Sciences, Department of Rehabilitation Sciences, Ghent University, Ghent, Belgium

4. Pain in Motion International Research Consortium, www.paininmotion.be

5. Department of Internal Medicine, Division of Rheumatology, Rush University Medical Center, Chicago, Illinois, USA

### Supplementary information

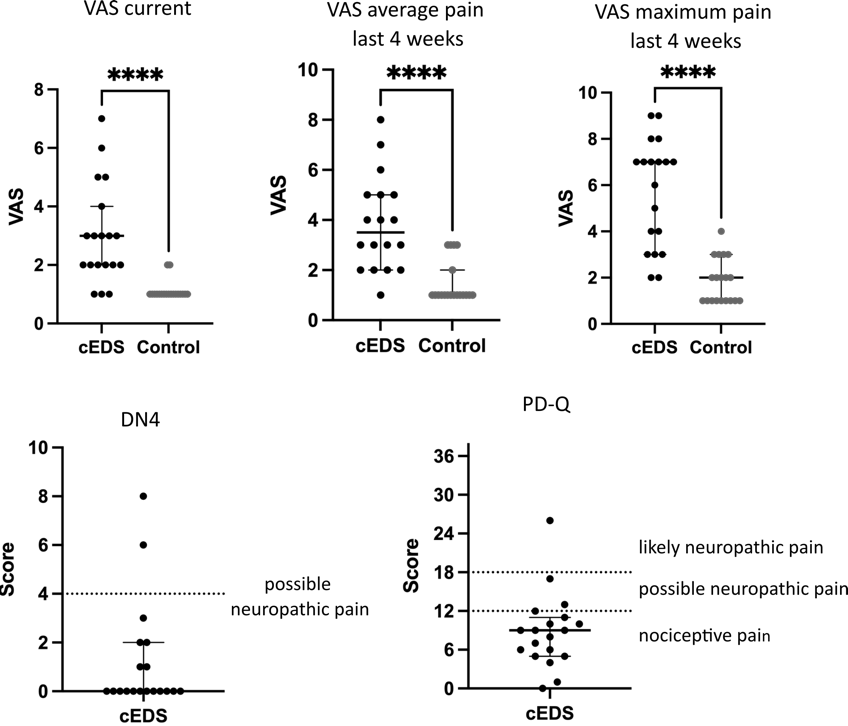

**Sup. Fig. 1: Pain severity and neuropathic pain**

VAS scores were used to assess subjective pain intensity on the day of the experimental procedures (current) and the average and maximum pain of the 4 weeks prior to the pain testing. In the cEDS group, symptoms of possible neuropathic pain were screened with the DN4 questionnaire where a score>4 indicates possible neuropathic pain, and the PD-Q (Pain-Detect questionnaire) where a score <12 indicates nociceptive pain, between 12-18 possible neuropathic pain and >12 likely neuropathic pain.

Normally distributed continuous variables are presented with ‘mean ± standard deviation, non-normally distributed continuous variables as ‘median (Interquartile range)’.

**** p<0.0001

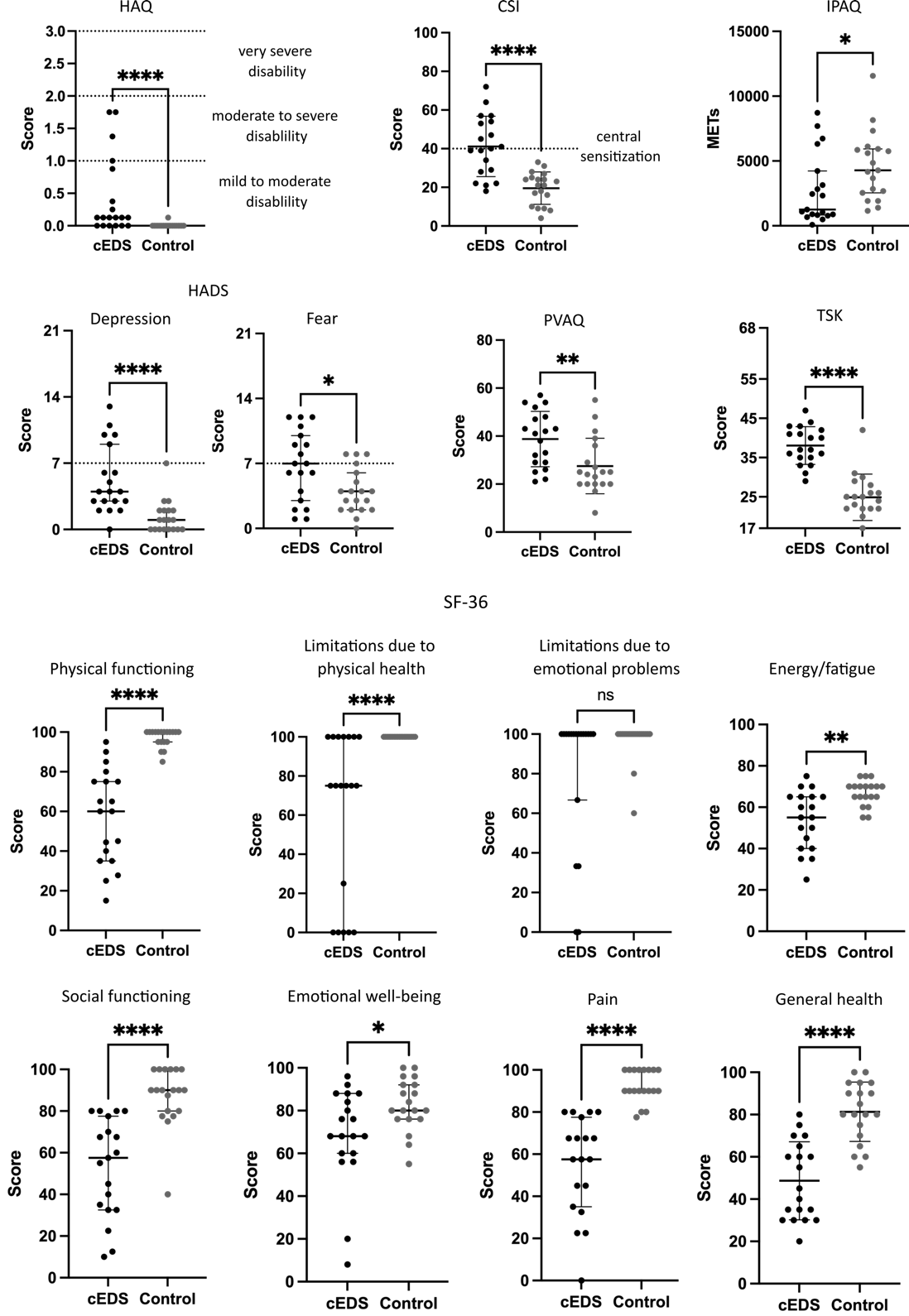

**Sup. Fig. 2: questionnaires**

Results of the HAQ (Health Assessment Questionnaire) with scores between 0-3 (0-1 mild to moderate disability, 1-2 moderate to severe disability, 2-3 severe disability), the CSI (Central Sensitization Inventory, score range 0-100) with scores >40 indicating possible symptoms of central sensitization, the IPAQ (International Physical Activity Questionnaire) scoring the metabolic equivalent task per week (METs), the HAD (Hospital Anxiety and Depression Scale, score range 0-21) with scores >7 denoting depression or anxiety on the respective subscales, the PVAQ (pain Vigilance and Awareness Questionnaire, score range 0-80) and the SF-36 (Short-Form 36 health survey) with its eight subscales (score range 0-100). Ns: non-significant, * p<0.05, ** p<0.01, **** p<0.0001
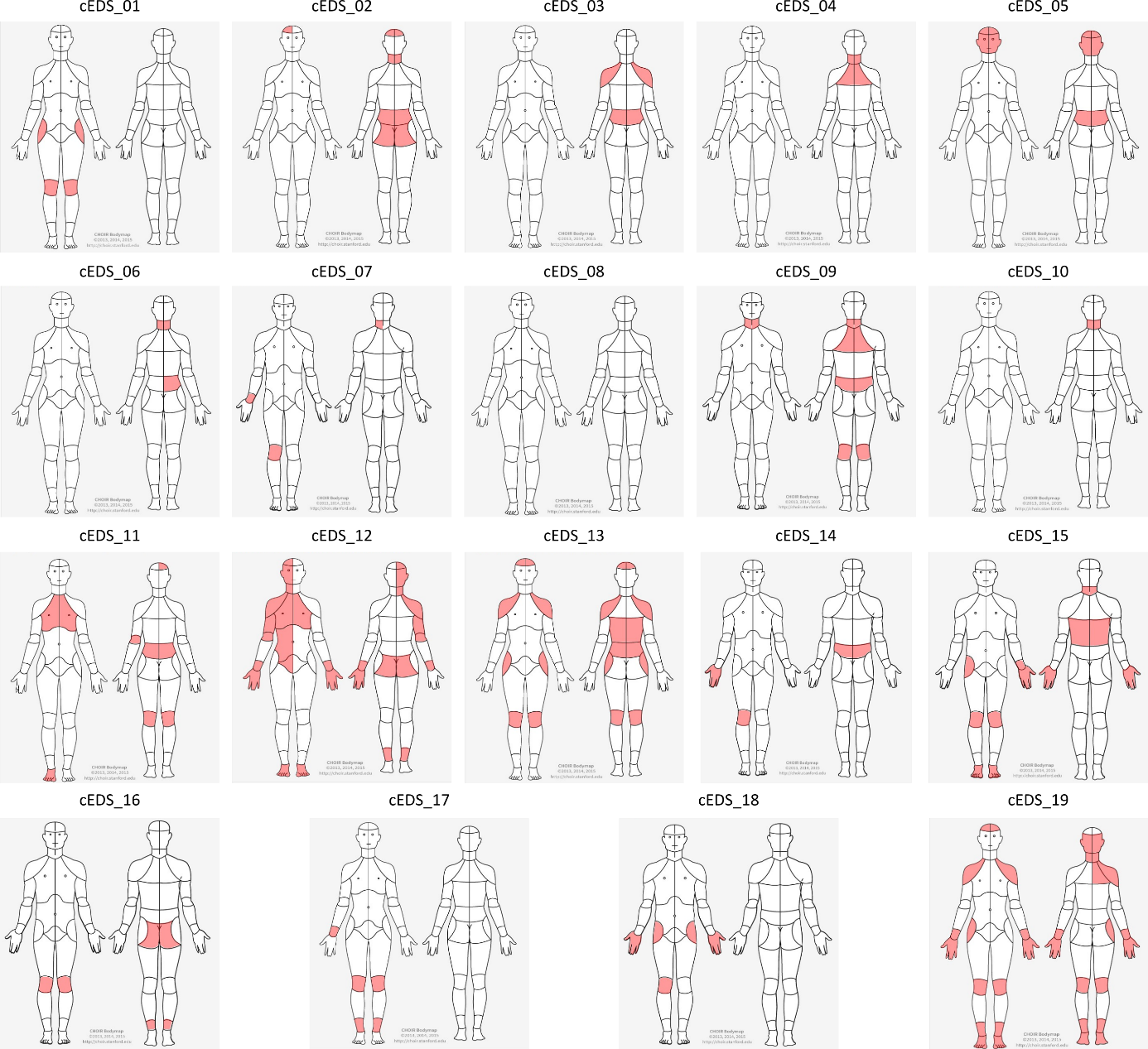

**Sup. Fig. 3: locations of regular pain on topographical body charts**

Participants were asked to indicate all body locations where they experienced regular pain the last 4 weeks. Regular pain was defined as pain that was present >24 hours the last 4 weeks.

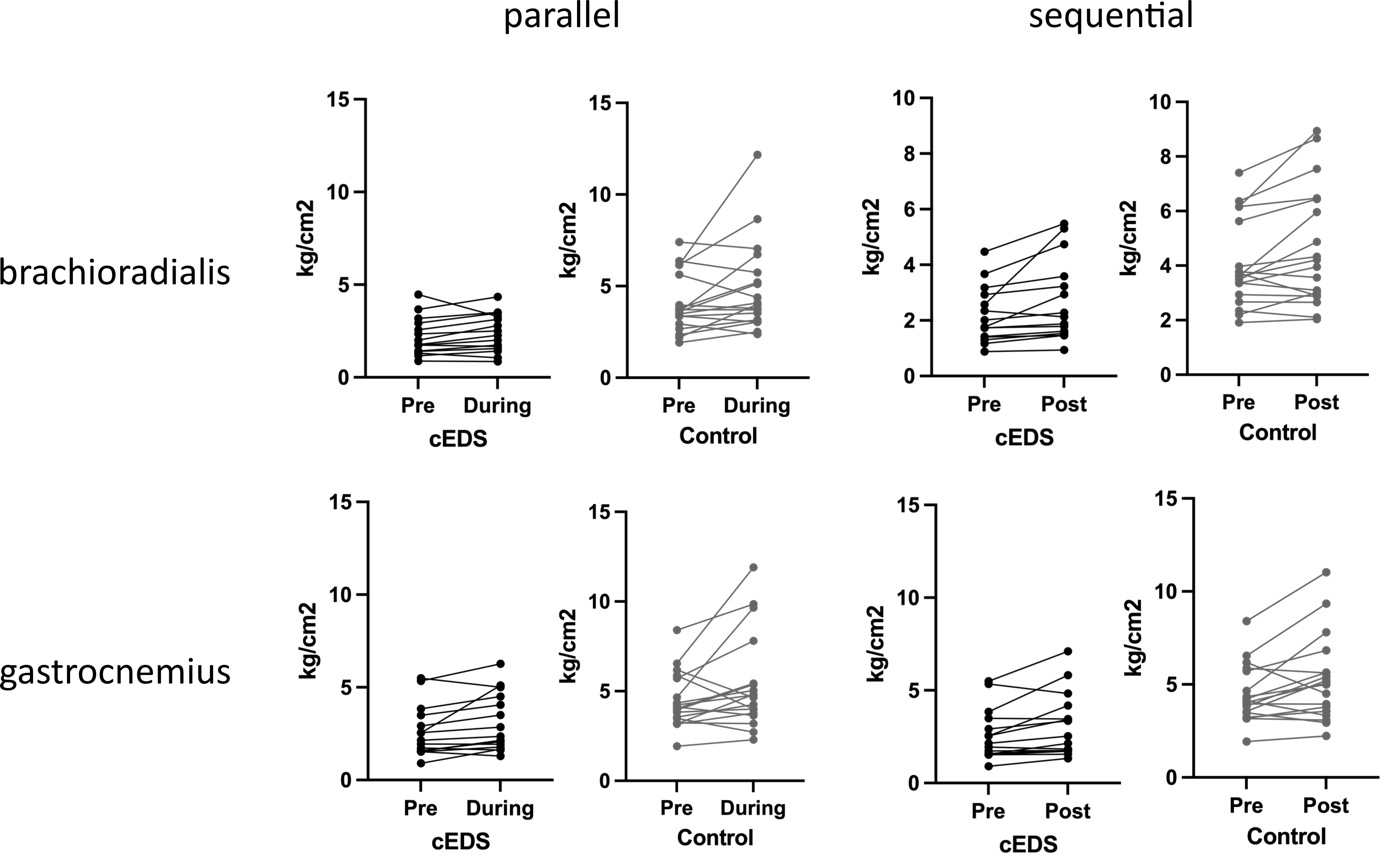

**Sup. Fig. 4**

CPM effects of all participants who finished the complete CPM protocol (both parallel and sequential methods).

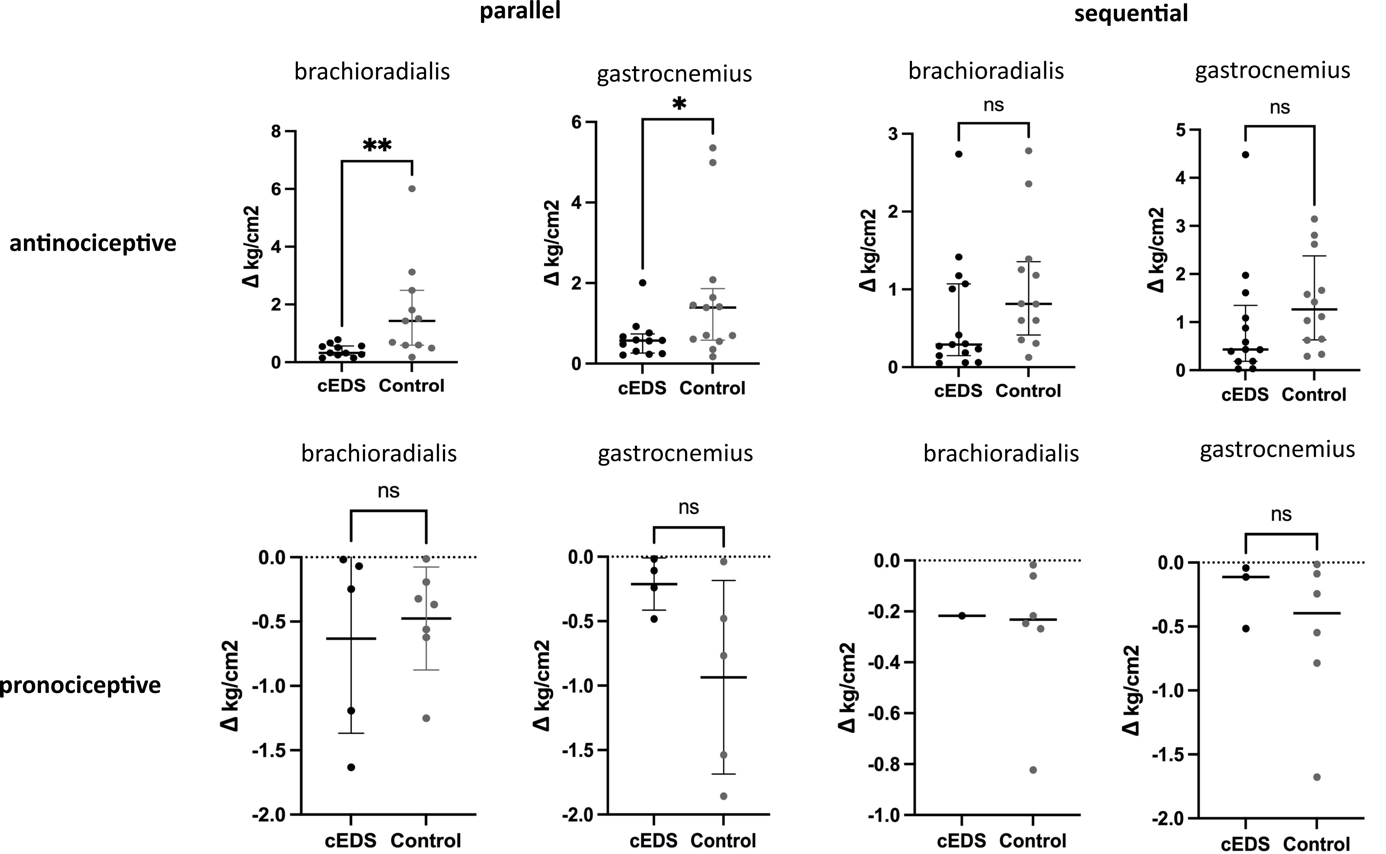

**Sup. Fig. 5**

anti- and pronociceptive CPM effects of the sequential and parallel CPM methods at both test locations. ns: non-significant, * p<0.05, ** p<0.01

***Sup. Table 1****: genetic characteristics of the included individuals with cEDS*

| Individual | Gene | Pathogenic defect | EDS variant database ID |
| --- | --- | --- | --- |
| cEDS_01 | *COL5A1* | c.5185del, p.(Thr1729Profs*) | COL5A1_000511 |
| cEDS_02 | *COL1A1* | c.934C>T, p.(Arg132Cys) | COL1A1_000043 |
| cEDS_03 | *COL5A1* | c.3184C>T, p.(Arg1062*) | COL5A1_00028 |
| cEDS_04 | *COL5A1* | c.5185del, p.(Thr1729Profs*) | COL5A1_000511 |
| cEDS_05 | *COL5A1* | Null allele^$^ |  |
| cEDS_06 | *COL5A1* | Null allele^$^ |  |
| cEDS_07 | *COL5A1* | c.4050dup, p.(Gly1351Argfs*) | COL5A1_000136 |
| cEDS_08 | *COL5A1* | c.4552C>T, p.(Gln1518*) | COL5A1_000139 |
| cEDS_09 | *COL5A1* | c.2034+1G>A | COL5A1_000101 |
| cEDS_10 | *COL5A1* | c.4916G>A, p.(Cys1639Tyr) | COL5A1_000141 |
| cEDS_11 | *COL5A1* | c.3258+33_3366+689delins, p.(Gly1087_Pro1122del) | COL5A1_000131 |
| cEDS_12 | *COL5A1* | c.4234G>T, p.(Gly854*) | COL5A1_00013 |
| cEDS_13 | *COL5A1* | c.3110del, p.(Thr1037Argfs*) | COL5A1_00024 |
| cEDS_14 | *COL5A1* | c.3110del, p.(Thr1037Argfs*) | COL5A1_00024 |
| cEDS_15 | *COL5A1* | c.3877C>T, p.(Glu1292Thrfs*) | COL5A1_000145 |
| cEDS_16 | *COL5A1* | c.74T>C, p.(Leu25Pro) | COL5A1_00051 |
| cEDS_17 | *COL5A1* | c.74T>G, p.(Leu25Arg) | COL5A1_00050 |
| cEDS_18 | *COL5A1* | c.74T>G, p.(Leu25Arg) | COL5A1_00050 |
| cEDS_19 | *COL5A2* | c.2194G>A, p.(Gly732Arg) | COL5A2_000175 |

$: a nonfunctional *COL5A1* allele was found, but the causal variant has not been identified.

***Sup. Table 2****: conditioned pain modulation*

|  |  |  |  | cEDS  n= 16^a^ | Control  n=18^a^ | *p*-value |
| --- | --- | --- | --- | --- | --- | --- |
| MPT (kg/cm^2^) | Parallel | brachioradialis |  | 2.27 (1.73) | 4.05 (2.86) | **≤0.001** |
|  |  | gastrocnemius |  | 2.35 (2.73) | 4.71 (2.28) | **0.005** |
|  | Sequential | brachioradialis |  | 2.13 (2.05) | 4.08 (3.55) | **0.005** |
|  |  | gastrocnemius |  | 3.12 ± 1.73 | 5.24 ± 2.30 | **0.004** |
| Antinociceptive CPM effect | Parallel | brachioradialis |  | 68.75% (n=11) | 61.11% (n=11) | 0.73 |
|  |  | gastrocnemius |  | 75.00% (n=12) | 72.22% (n=13) | >0.999 |
|  | Sequential | brachioradialis |  | 93.75% (n=14) | 66.67% (n=12) | 0.48 |
|  |  | gastrocnemius |  | 81.25% (n=13) | 66.67% (n=12) | 0.45 |
| CPM effect | Parallel | brachioradialis | Δ | 0.26 (0.58) | 0.54 (1.92) | 0.31 |
|  |  |  | % | 9.07 ± 18.17 | 0.23 ± 23.16 | 0.17 |
|  |  | gastrocnemius | Δ | 0.39 (0.68) | 0.65 (1.65) | 0.38 |
|  |  |  | % | 16.10 (31) | 17.77 (40) | 0.82 |
|  | Sequential | brachioradialis | Δ | 0.28 (0.97) | 0.48 (1.30) | 0.99 |
|  |  |  | % | 13.27 (18) | 11.62 (26) | 0.48 |
|  |  | gastrocnemius | Δ | 0.41 (1.40) | 0.64 (1.73) | 0.88 |
|  |  |  | % | 20.02 ± 19.76 | 16.59 ± 25.16 | 0.66 |
| VAS | 2 minutes | |  | 4.56 ± 1.25 | 3.9 ± 1.72 | 0.11 |
|  | 6 minutes | |  | 5.64 ± 2.13 | 4.12 ± 2.48 | 0.08 |

Normally distributed continuous variables are presented with ‘mean ± standard deviation’, non-normally distributed continuous variables as ‘median (Interquartile range)’.

^a^ Participants who completed the CPM protocol; Δ absolute differences in MPT expressed as kg/cm^2^, % relative difference in MPT expressed as percentage.
